# Genetically proxied NLRP3 activity and coronary artery disease risk: a drug-target Mendelian randomization study

**DOI:** 10.64898/2026.11.20.26368563

**Authors:** Nick Hirschmüller, Paul Carter, Stephen Burgess, Elias Allara, James E. Peters, Liam Gaziano, Rima Mustafa, Murray C. H. Clarke, Ziad Mallat, Dirk S. Paul, Adam S. Butterworth

## Abstract

**Background and Aims:** Preclinical studies implicate NLRP3 inflammasome activation, which drives release of interleukin-1β and interleukin-18, in vascular inflammation and atherosclerosis, and selective NLRP3 inhibitors are now in clinical development. However, whether lowering NLRP3 activity reduces coronary artery disease (CAD) risk in humans is unknown. We used human genetics to estimate this effect and its mediating pathways.

**Methods:** A genetic NLRP3 activity score was derived by combining the effects of eight *cis*-*NLRP3* variants on *NLRP3* expression and three inflammatory biomarkers: C-reactive protein, glycoprotein acetylation, and neutrophil count. The score was validated against circulating interleukin-1β and interleukin-18. Drug-target Mendelian randomization was used to estimate its association with CAD, cardiometabolic traits and other diseases studied in NLRP3 inhibitor trials. Mediation by cardiometabolic risk factors was quantified using the product-of-coefficients method.

**Results:** A lower genetically proxied NLRP3 activity score was associated with lower circulating interleukin-1β and interleukin-18, but with higher CAD risk (OR = 1.21, 95% CI 1.02–1.45, *P* = 0.032). It was also associated with higher systolic blood pressure, apolipoprotein B levels, and type 2 diabetes risk, which together mediated an estimated 87% of the association with CAD. The residual direct association with CAD was attenuated (OR = 1.03, 95% CI 0.85–1.24, *P* = 0.79). In contrast, a lower score was nominally associated with lower risk of gout, pericarditis, and Parkinson’s disease.

**Conclusions:** Lifelong lower genetically proxied NLRP3 activity was associated with higher CAD risk, largely mediated by adverse cardiometabolic effects, with no evidence of protection after accounting for them. These findings do not provide genetic support for lowering NLRP3 activity to reduce CAD risk.

## Introduction

Despite optimal management of established cardiovascular risk factors, including low-density lipoprotein cholesterol (LDL-C) and blood pressure, patients with established coronary artery disease (CAD) remain at considerable risk of recurrent cardiovascular events. Persistent vascular inflammation, reflected by elevated high-sensitivity C-reactive protein (hsCRP) levels, is recognized as a major contributor to this residual cardiovascular risk^1^. The CANTOS trial provided clinical proof of concept that selective inhibition of interleukin-1β (IL-1β), a pro-inflammatory cytokine, reduces recurrent cardiovascular events independently of lipid lowering^2^. Trials of the broad anti-inflammatory drug colchicine have yielded mixed results. Colchicine reduced cardiovascular events after recent myocardial infarction in COLCOT^3^ and among patients with chronic coronary disease in LoDoCo2^4^, but not after acute myocardial infarction in CLEAR-SYNERGY^5^ or after ischaemic stroke in CONVINCE^6^ and CHANCE-3^7^. Recently, the phase 3 ZEUS trial found no reduction in cardiovascular events with interleukin-6 (IL-6) inhibition in patients with atherosclerotic disease, chronic kidney disease, and elevated hsCRP^8^. Together, these findings suggest that cardiovascular benefit depends on both the inflammatory pathway targeted and the clinical context, rather than on inflammation lowering per se.

One inflammatory pathway that has gained considerable attention is centred on NLRP3. NLRP3 is the sensor protein of the NLRP3 inflammasome, a multiprotein complex comprising NLRP3, the adaptor ASC, and caspase-1, whose activation promotes the maturation and release of IL-1β and interleukin-18 (IL-18)^9^. These cytokines promote downstream inflammatory signalling pathways, including IL-6 signalling, which drives the acute-phase response characterized by elevated inflammatory markers such as C-reactive protein (CRP) and glycoprotein acetylation (GlycA), a composite measure of glycosylated acute-phase proteins^10,11^. Concurrently, NLRP3 inflammasome activation increases the production of circulating myeloid cells, predominantly neutrophils^12^.

In preclinical models of atherosclerosis, genetic deficiency or pharmacological inhibition of NLRP3 has been reported to reduce vascular inflammation and plaque burden ^13,14^. Together with the evidence linking NLRP3 inflammasome-related cytokines to vascular inflammation, these findings have motivated the development of small-molecule inhibitors that act directly on the NLRP3 sensor, several of which have entered clinical testing^15^. However, no phase 3 trial has yet established whether selective NLRP3 inhibition reduces cardiovascular events in humans.

In the absence of large randomized clinical trials, drug-target Mendelian randomization (MR) can provide genetic evidence on whether a target causally influences disease risk, using naturally occurring variation near drug-target genes to proxy differences in target activity. This approach has precedent in cardiovascular drug development. For example, genetic variation in *PCSK9* supported LDL-C lowering through PCSK9 inhibition as a strategy expected to reduce cardiovascular risk^16^. Conversely, genetic proxies of lower lipoprotein-associated phospholipase A_2_ (Lp-PLA_2_) activity were not associated with lower coronary heart disease risk, consistent with the lack of benefit observed for pharmacological Lp-PLA_2_ inhibition^17^.

Human genetic evidence on NLRP3 is limited. A study based on a single variant associated with *NLRP3* expression in blood linked higher NLRP3 activity to higher cardiovascular mortality^18^. In this study, we combined the effects of multiple variants on *NLRP3* expression with their effects on downstream inflammatory biomarkers into an NLRP3 activity score, validated it against IL-1β and IL-18, and used drug-target MR to test its association with CAD, the pathways involved, and diseases for which NLRP3 inhibitors have been, or are being, evaluated.

## Methods

### Study design

We used drug-target MR to test whether genetically proxied NLRP3 activity, represented by an NLRP3 activity score, was associated with CAD risk (Figure 1). We constructed the score from eight *cis*-acting variants at the *NLRP3* locus, using their independent associations with *NLRP3* expression and well-powered circulating inflammatory biomarkers downstream of NLRP3 inflammasome activation (Supplementary Information). We validated this score against canonical NLRP3 inflammasome effector cytokines IL-1β and IL-18 before estimating its association with CAD. We further assessed mediation by cardiometabolic risk factors and explored additional diseases for which selective NLRP3 inhibitors have been or are being evaluated in clinical trials.

**Figure 1.**
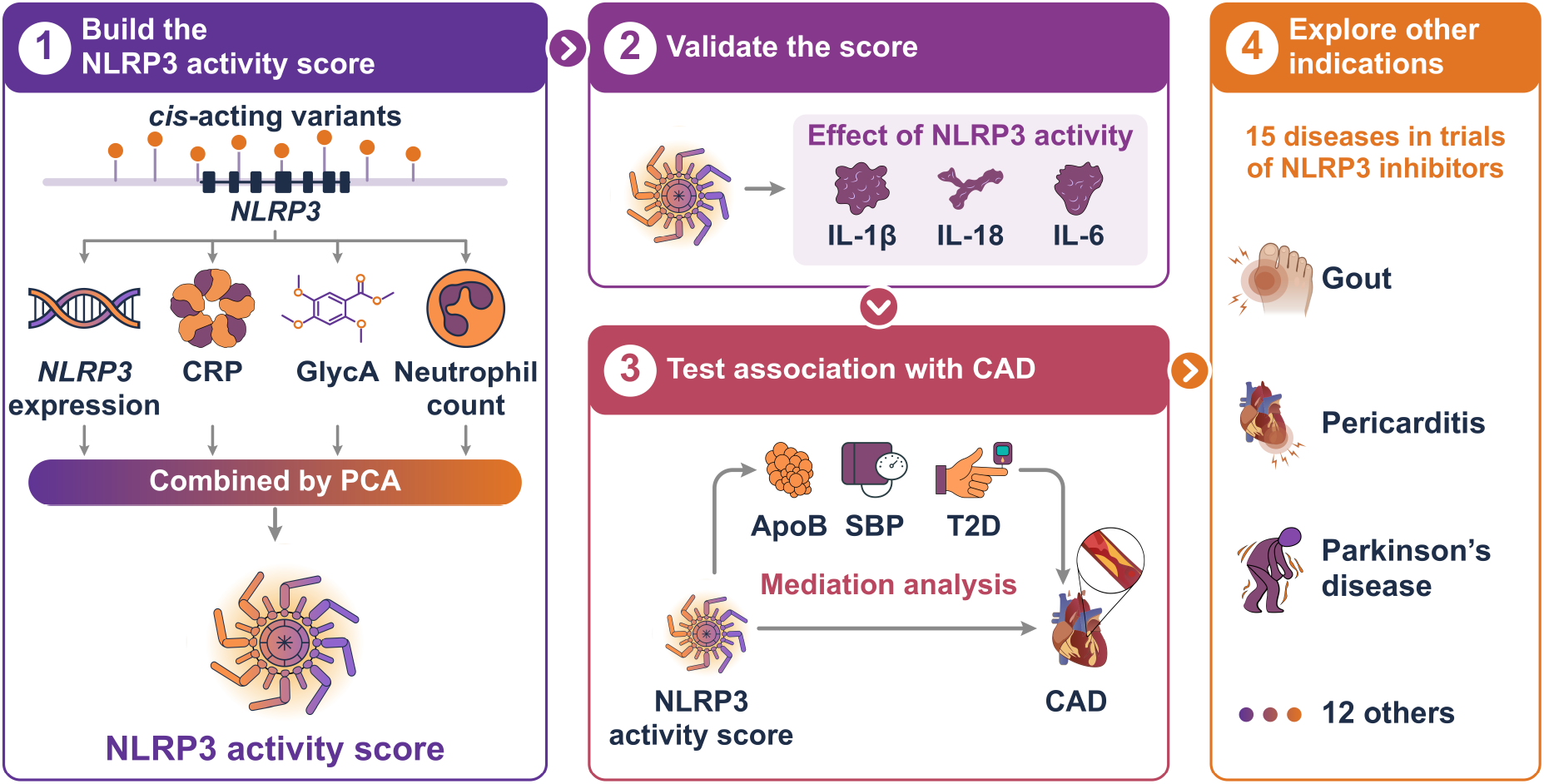
Study design. The NLRP3 activity score was constructed from *cis*-acting variants at the *NLRP3* locus associated with *NLRP3* expression, CRP, GlycA, and neutrophil count, whose effect sizes were combined by PCA. The score was validated against the canonical inflammasome effectors IL-1β and IL-18. Using Mendelian randomization, the score was applied to estimate the effect of genetically proxied NLRP3 activity on CAD risk and, through mediation analysis, to quantify the extent to which this effect was mediated by cardiometabolic risk factors, namely ApoB, SBP, and T2D. The score was further applied across additional clinical indications, including gout, pericarditis, and Parkinson’s disease.

### Data sources

Across analyses, we prioritized large publicly available genome-wide association study (GWAS) summary statistics from European-ancestry participants to maximize statistical power while maintaining ancestry consistency with the instrument-construction datasets. Analyses included men and women combined, because sex-specific summary statistics were not available for most datasets and would have reduced statistical power.

### Datasets for instrument construction and validation

Instrument construction used GWAS summary statistics for circulating CRP levels, GlycA concentrations, neutrophil count, and whole-blood *NLRP3* expression. Summary statistics for CRP levels were obtained from a UK Biobank and CHARGE meta-analysis including 575,531 European-ancestry participants^19^. Summary statistics for GlycA concentrations were obtained from 434,646 European-ancestry UK Biobank participants, with GlycA quantified using the Nightingale Health nuclear magnetic resonance metabolomics platform^20^. Neutrophil count summary statistics were obtained from 519,288 European-ancestry participants of a multi-ancestry blood-cell trait meta-analysis^21^. Whole-blood *NLRP3* expression quantitative trait locus data were obtained from 4,732 European-ancestry INTERVAL participants, with expression measured by RNA sequencing^22^. For instrument validation, we used plasma protein GWAS summary statistics from the full UK Biobank Pharma Proteomics Project cohort, comprising up to 51,637 predominantly European-ancestry participants per protein profiled using the Olink Explore platform, to maximize statistical power and variant coverage^23^. IL-1β and IL-18 were available only in this smaller proteomic dataset and were therefore used to validate the score rather than to construct it. Further details are provided in Supplementary Table 1.

### Datasets for disease outcomes

The primary disease outcome was CAD, for which we combined four European-ancestry datasets: a CAD GWAS meta-analysis including 181,522 cases and 984,168 controls^24^, coronary atherosclerosis in the Million Veteran Program (MVP), including 124,302 cases and 300,039 controls^25^, coronary atherosclerosis in All of Us, including 19,823 cases and 150,340 controls^26^, and coronary atherosclerosis in FinnGen (release 12), including 63,307 cases and 416,171 controls^27^. Variant–CAD association estimates were combined by fixed-effect inverse-variance weighted meta-analysis, and the resulting meta-analysed summary statistics served as the primary CAD outcome for MR. As a measure of subclinical atherosclerosis, we obtained summary statistics for imaging-defined atherosclerosis from the Swedish CArdioPulmonary bioImage Study (SCAPIS), comprising coronary artery calcification (n = 26,000), coronary plaque burden quantified as the segment involvement score on CT angiography (n = 24,811), and ultrasound-assessed carotid plaque (n = 26,807), which have no sample overlap with the datasets used for instrument construction or with the CAD datasets^28^. Further details are provided in Supplementary Table 2. We additionally obtained European-ancestry summary statistics for 15 diseases for which selective NLRP3 inhibitors have been, or are being, evaluated in clinical trials. Six were taken from MVP^25^, and the others from GWAS of amyotrophic lateral sclerosis^29^, asthma^30^, COVID-19^31^, knee osteoarthritis^32^, Parkinson’s disease^33^, pericarditis^34^, rheumatoid arthritis^35^, type 2 diabetes^36^, and ulcerative colitis^37^. Further details are provided in Supplementary Table 3.

### Datasets for cardiometabolic and lifestyle traits

We used publicly available European-ancestry GWAS summary statistics for cardiometabolic and lifestyle traits relevant to CAD risk, including lipid traits^20,38,39^, blood pressure^40^, body mass index^41^, type 2 diabetes^36^, smoking^42^, and alcohol consumption^42^. These traits were screened for association with the NLRP3 activity score, and associated traits (*P* < 0.05) were considered as candidate mediators. Further details are provided in Supplementary Table 4.

### Genetic instrument construction

To construct a genetic proxy for NLRP3 activity, we used whole-blood *NLRP3* expression and three well-powered inflammatory traits that are elevated downstream of NLRP3 inflammasome activation and have strong statistical evidence of genetic association at the *NLRP3* locus: circulating CRP and GlycA levels, and blood neutrophil count (Supplementary Information). For each trait, variants within ±150 kb of *NLRP3* were clumped for linkage disequilibrium (LD) at *r*² < 0.1 using 11,863 European-ancestry whole-genome sequences from INTERVAL as the LD reference panel. We merged the clumped signals across traits and retained variants associated at genome-wide significance ( *P* < 5×10⁻⁸) with at least two traits, yielding eight variants for instrument construction (Supplementary Figure 1). To combine the four traits into a single exposure, we applied principal component analysis (PCA) to the centred and scaled trait-specific genetic effect estimates and used the first principal component loadings as data-driven trait weights. The resulting NLRP3 activity score was scaled to CRP, such that a one-unit decrease corresponds approximately to a one standard deviation decrease in CRP. Standard errors were estimated using the delta method^43^. Instrument strength was summarized by the F-statistic for the eight variants jointly, while accounting for their residual LD^44^. As a methodological positive control, we applied the same procedure at the *IL1RN* locus to construct a genetically proxied interleukin-1 receptor antagonist (IL1Ra) activity score and tested it against plasma IL1Ra concentration and against gout and rheumatoid arthritis.

### Colocalization analyses

To evaluate whether inflammatory biomarker associations at the *NLRP3* locus share a causal variant, we performed multi-trait colocalization using HyPrColoc^45^ (version 0.0.2) across whole-blood *NLRP3* expression, circulating CRP levels, GlycA concentrations, and blood neutrophil count, using variants within ±200 kb of *NLRP3*. To isolate the association cluster within the *NLRP3* gene from the stronger peak upstream of it, we conditioned the *NLRP3* expression summary statistics on that peak’s lead variant (rs6689545) using GCTA-COJO^46^ (version 1.94.1) with the INTERVAL reference panel. The conditioned expression and unconditioned biomarker summary statistics were then used for colocalization. HyPrColoc was run with default variant-specific priors (prior.1 = 1×10^-4^, prior.c = 0.02), and a posterior probability > 0.8 that the traits share a causal variant was interpreted as strong evidence of colocalization. To test for colocalizing signals elsewhere in the region, we repeated HyPrColoc, with the same inputs and priors, in the flanking regions from 200 kb to 1 Mb upstream and downstream of *NLRP3*.

### Mendelian randomization analyses

We used two-sample MR to estimate associations of the NLRP3 activity score with circulating biomarkers, cardiometabolic traits, and disease outcomes. All estimates were oriented to represent lower genetically proxied NLRP3 activity, mirroring pharmacological inhibition. The primary estimator was random-effects inverse-variance weighted (IVW) MR. Weighted median MR was used as a robustness analysis and the MR-Egger intercept was used to test whether variant effects on the outcome were proportional to their effects on the exposure. MR-Egger was not used as a causal estimator, as its instrument strength independent of direct effect (InSIDE) assumption is not well suited to variants within a single gene region^47^. IVW and MR-Egger accounted for residual LD using the INTERVAL reference panel. An instrument variant missing from an outcome dataset was replaced by the variant in highest LD with it in the same panel (*r*² ≥ 0.9 within 50 kb), where one was available. For CAD, we additionally performed leave-one-out analyses, and repeated instrument selection and score construction at relaxed LD clumping thresholds (*r*² 0.2–0.6). We also repeated the biomarker, cytokine and CAD analyses using a conventional instrument based on *NLRP3* expression alone (Supplementary Information). Estimates are reported as betas for continuous outcomes, odds ratios (ORs) for binary outcomes, and proportional odds ratios for ordinal outcomes, with 95% confidence intervals (CIs). Analyses used the MendelianRandomization^44^ (version 0.10.0) and MVMR^48^ (version 0.4.2) R packages.

### Evaluation of the NLRP3 activity score

We first assessed internal consistency by estimating associations of a lower NLRP3 activity score with the downstream inflammatory biomarkers used for score construction: circulating CRP levels, GlycA concentrations, and blood neutrophil count. We then evaluated circulating IL-1β, IL-18, and IL-6 levels as external biological readouts. IL-1β and IL-18 were selected because they are canonical cytokine products of NLRP3 inflammasome activation, while IL-6 was included as a downstream cytokine linking interleukin-1 signalling to the acute-phase response^9–11^.

### Mediation analysis

To investigate whether the association between a lower genetically proxied NLRP3 activity score and higher CAD risk could be explained by established cardiovascular risk factors, we performed mediation analyses for systolic blood pressure (SBP), apolipoprotein B (ApoB), and type 2 diabetes (T2D). These traits were associated with the NLRP3 activity score in the cardiometabolic and lifestyle trait screen and are established risk factors for CAD^49^. ApoB was chosen as the single lipid mediator^50^, and SBP was preferred to the correlated diastolic blood pressure (Supplementary Information). We used the product-of-coefficients method to estimate indirect effects^51^, multiplying the MR effect of the score on each mediator by the mutually adjusted effect of that mediator on CAD from multivariable MR (MVMR) ^48^. MVMR used genome-wide significant variants for SBP, ApoB and T2D from their source GWAS^20,36,40^, and instrument strength was assessed with the conditional F-statistic^48^ (Supplementary Information). All CAD effects came from the meta-analysis of the four CAD datasets, and all mediation analyses used the log-odds scale. The joint indirect effect was the sum of the three indirect effects, the proportion mediated was the joint indirect effect divided by the total effect, and the residual direct effect was the total effect minus the joint indirect effect. Uncertainty in the joint indirect effect was estimated with the multivariate delta method^52^, and the 95% CI of the proportion mediated by Monte Carlo simulation.

### *NLRP3* predicted loss-of-function analysis

To test whether rare *NLRP3* predicted loss-of-function (pLOF) variants were associated with circulating inflammatory readouts, we performed gene-based burden tests for CRP levels, GlycA concentrations, and blood neutrophil count among UK Biobank participants with whole exome sequencing (WES) data and available biomarker measurements. Burden tests were performed using SAIGE-GENE+ (version 1.5.0)^53^. Effect estimates are per unit of the weighted burden score. pLOF variants were defined as frameshift, stop-gained, splice donor, or splice acceptor variants using variant consequences reported in gnomAD (version 4.1)^54^. Analyses were restricted to individuals of genetically inferred European ancestry^55^. Analogous pLOF burden analyses were performed for all protein-coding genes within ±110 kb of *NLRP3*. Phenotype processing is described in the Supplementary Information.

### *NLRP3* gain-of-function analysis

Pathogenic gain-of-function variants in *NLRP3* were obtained from the INFEVERS registry^56^ and Cosson *et al*.^57^. Carriers were identified among UK Biobank participants with WES data and available biomarker measurements. Only one of these gain-of-function variants was observed in UK Biobank, corresponding to GRCh38 chr1:247424765 C>T (p.Ala439Val). All three carriers were heterozygous. Due to the limited number of carriers, CRP levels, GlycA concentrations, and blood neutrophil counts were reported as percentiles relative to non-carriers. Phenotype processing is described in the Supplementary Information.

### Proteome-wide association and over-representation analyses

To characterize broader circulating protein changes associated with lower genetically proxied NLRP3 activity, we performed two-sample MR using plasma protein GWAS summary statistics from the UK Biobank Pharma Proteomics Project^23^. Associations were estimated using IVW MR with LD correction, as described above. Proteins were considered significant using a Bonferroni correction based on the effective number of independent protein tests^58^, yielding a threshold of *P* < 2.7×10^-5^. Significant proteins were tested for enrichment of Gene Ontology Biological Process terms separately by direction of association. Details of the effective test correction and over-representation analysis are provided in the Supplementary Information.

### Exploratory analyses across diseases under investigation for NLRP3 inhibition

Disease outcomes for exploratory MR analyses were selected based on diseases for which selective NLRP3 inhibitors have been, or are being, evaluated in registered clinical trials, as identified from a structured search of four clinical trial registries conducted on 28 April 2026 (see Supplementary Information for full details). Identified diseases were included where publicly available GWAS summary statistics of sufficient sample size were available. The included exploratory outcomes and corresponding GWAS sources are provided in Supplementary Table 3. As these analyses were exploratory, nominal P-values are reported without adjustment for multiple testing.

### Software and statistical significance

Analyses were performed in R (version 4.3.1). Colocalization used HyPrColoc (version 0.0.2), conditional analysis used GCTA-COJO (version 1.94.1), and LD clumping used PLINK 2 (version 2.00a6). All P-values are two-sided, and P < 0.05 was considered statistically significant unless stated otherwise.

## Results

### Genetic associations with inflammatory biomarkers at the *NLRP3* locus

Using publicly available GWAS summary statistics (Supplementary Table 1), we identified associations of variants at the *NLRP3* locus with whole-blood *NLRP3* expression (rs58546652, *P* = 1.04×10^-58^, *n* = 4,732), circulating CRP levels (rs12239046, *P* = 1.34×10^-87^, *n* = 575,531), GlycA concentrations (rs56188865, *P* = 4.7×10^-28^, *n* = 434,646), and blood neutrophil count (rs56188865, *P* = 1.93×10^-43^, *n* = 519,288) (Figure 2A). Multi-trait colocalization across *NLRP3* expression and the three circulating inflammatory biomarkers showed strong evidence of a shared causal variant between the four traits (posterior probability = 0.97). No additional colocalizing signals were observed in the flanking regions between 200 kb and 1 Mb on either side of *NLRP3* (posterior probability < 0.13), indicating that the inflammatory biomarker associations shared across traits in this region are centred on *NLRP3* (Supplementary Figure 2). Together, these findings support a coherent NLRP3-linked inflammatory signal at the locus and identify variants suitable for genetic instrument construction.

**Figure 2.**
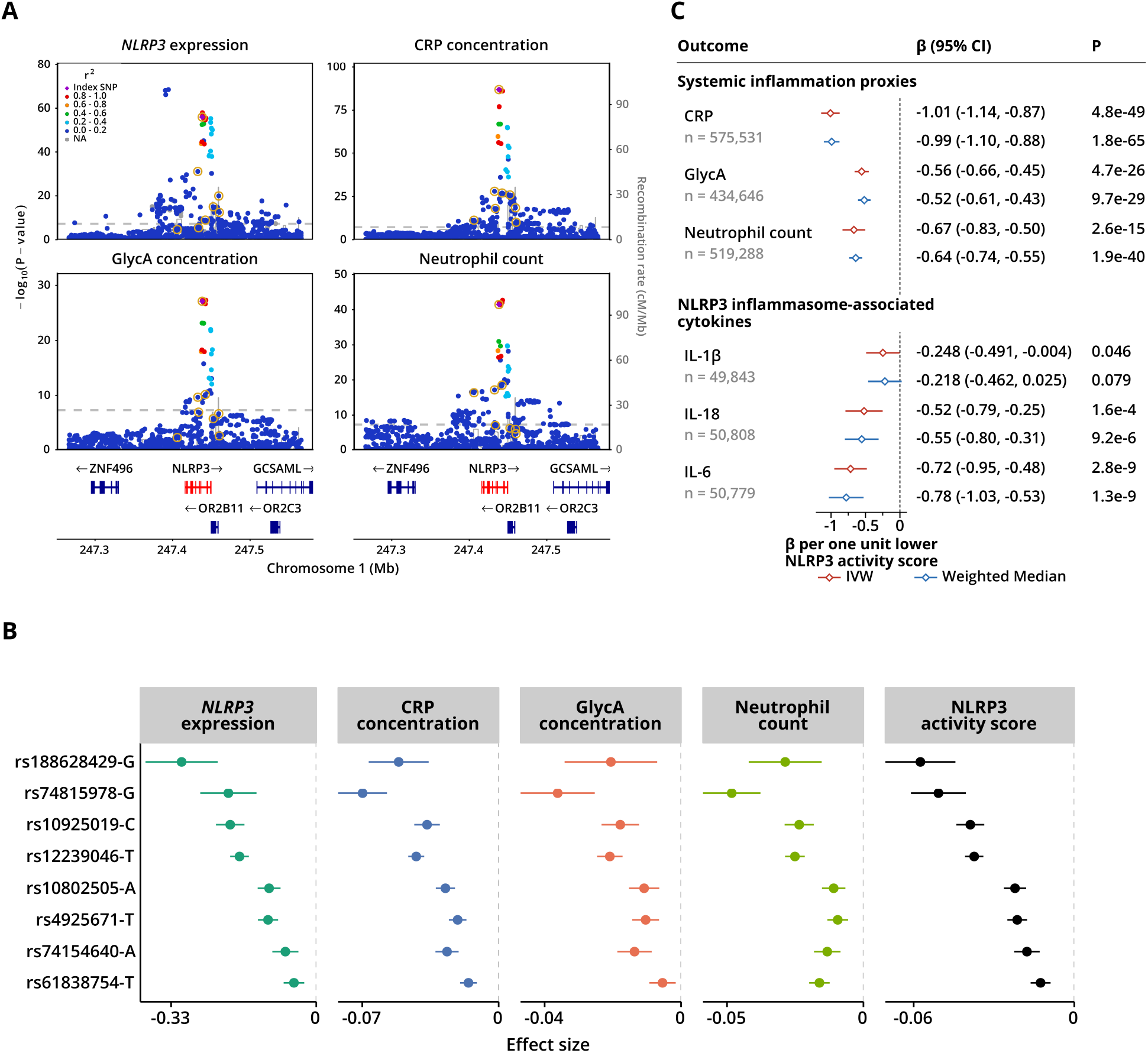
Construction and validation of the NLRP3 activity score. **(A)** Regional association plots for *NLRP3* expression, CRP concentration, GlycA concentration, and neutrophil count at the *NLRP3* locus. *NLRP3* expression is shown unconditioned. Its strongest association (rs6689545) is an independent upstream signal with little effect on CRP, GlycA, or neutrophil count, and was conditioned out before colocalization. Variants are coloured by linkage disequilibrium (*r*²) with rs12239046 (purple diamond). The eight variants taken forward as instruments are circled in gold. **(B)** Per-allele effect sizes and 95% confidence intervals of each variant on individual traits and on the NLRP3 activity score. The latter represents the PC1-weighted composite of effect sizes across the four traits. The effect allele, shown after each rsID, is the allele that corresponds to lower NLRP3 activity score. **(C)** Forest plot of MR estimates for the effect of the NLRP3 activity score on the inflammatory biomarkers used in its construction (CRP, GlycA, and neutrophil count) and on cytokines not used in its construction (IL-1β, IL-18, and IL-6). Estimates are reported as SD changes per one-unit lower NLRP3 activity score, from inverse-variance weighted and weighted median MR. PC1, first principal component.

### Genetic instrument selection and validation

Because neither *NLRP3* expression nor any single circulating biomarker specifically measures NLRP3 activity, we constructed a composite genetic exposure combining whole-blood *NLRP3* expression with downstream circulating inflammatory readouts (Supplementary Information). We identified genetic signals associated with at least two of the four traits (*P* < 5×10^-8^). From these signals, we selected eight approximately independent variants (*r*^2^ < 0.1, Supplementary Figure 1). We then applied PCA to their effect estimates across the four traits. The first principal component explained 96.8% of the variation across the traits’ genetic associations. We used the weights derived from this component to combine these associations into the NLRP3 activity score (Figure 2B, Supplementary Table 5), which served as the exposure in subsequent MR analyses. The eight variants explained 8.3% of the variance in *NLRP3* expression, 0.11% in CRP, 0.046% in GlycA, and 0.064% in neutrophil count, and the joint F-statistic for the score was 92.0, indicating a strong instrument.

To evaluate the NLRP3 activity score, we used two-sample MR to estimate its associations with downstream biomarkers. We first confirmed that a lower genetically proxied NLRP3 activity score was associated with lower CRP, GlycA, and neutrophil count, consistent with the score’s construction (Figure 2C). We then tested associations with NLRP3-related cytokines that were not used in score construction. A lower genetically proxied NLRP3 activity score was associated with lower circulating IL-1β (*β* = -0.25, 95% CI -0.49 to -0.004, *P* = 0.046), IL-18 (*β* = -0.52, 95% CI -0.79 to -0.25, *P* = 0.00016) and IL-6 (*β* = -0.72, 95% CI -0.95 to -0.48, *P* = 2.8×10^-9^). Effect estimates were directionally consistent across IVW and weighted median estimators (Supplementary Table 6). Together, these findings support the NLRP3 activity score as capturing the expected downstream inflammatory consequences of lower NLRP3 activity.

### Association of the NLRP3 activity score with coronary artery disease risk

We performed two-sample MR to estimate the association between the genetically proxied NLRP3 activity score and CAD risk using a fixed-effect meta-analysis of four independent European-ancestry GWAS datasets, comprising 388,954 cases and 1,850,718 controls. Despite being associated with lower inflammatory biomarkers, a lower NLRP3 activity score was associated with higher CAD risk (OR = 1.21, 95% CI 1.02 to 1.45, *P* = 0.032) (Figure 3A). Estimates were directionally concordant across all four contributing datasets and were preserved under the weighted median estimator (OR = 1.23, 95% CI 1.06 to 1.42, *P* = 0.005). The MR-Egger intercept differed from zero (intercept = 0.009, 95% CI 0.002 to 0.016, *P* = 0.018), indicating that the variants’ effects on CAD are not fully proportional to their effect on the NLRP3 activity score (Supplementary Table 7).

**Figure 3.**
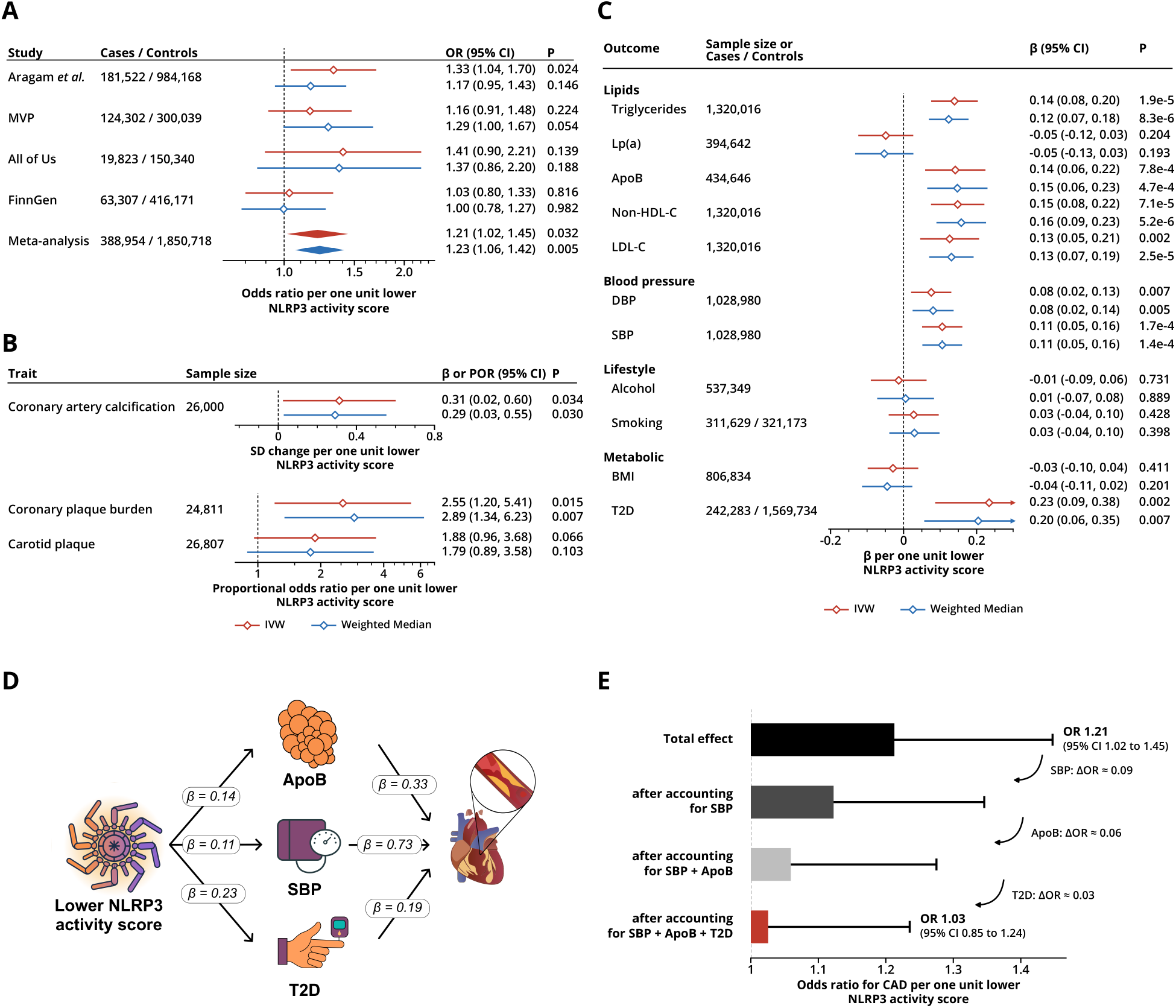
Association of the NLRP3 activity score with coronary artery disease risk and its mediation by cardiometabolic risk factors. **(A)** Forest plot of MR estimates for the effect of a lower NLRP3 activity score on CAD risk in each contributing GWAS (Aragam *et al.*, MVP, All of Us, and FinnGen) and in their fixed-effect meta-analysis. **(B)** MR estimates for the association of a lower NLRP3 activity score with coronary artery calcification, coronary plaque burden, and carotid plaque. Coronary artery calcification is reported as SD change, whereas coronary and carotid plaque outcomes are reported as proportional odds ratios. **(C)** Forest plot of MR estimates for the effect of a lower NLRP3 activity score on cardiometabolic risk factors, grouped into lipid, blood pressure, lifestyle, and metabolic traits. Estimates for continuous outcomes are reported in SD units and estimates for T2D as log odds ratios. **(D)** Mediation diagram of the pathways from a lower NLRP3 activity score to CAD through ApoB, SBP, and T2D, with each mediator-to-CAD effect estimated in mutually adjusted MVMR. Effects of the score on ApoB and SBP are in SD units and on T2D in log odds. Mediator-to-CAD effects are log odds ratios for CAD per SD higher ApoB or SBP, and per one-unit higher log odds of T2D liability. **(E)** Waterfall plot of the progressive attenuation of the total effect of a lower NLRP3 activity score on CAD risk as SBP, ApoB, and T2D are sequentially accounted for, resulting in the residual direct effect. Models were refitted at each step using the instrument set for the included mediators. ΔOR indicates the decrease in odds ratio between successive models. DBP, diastolic blood pressure; Lp(a), lipoprotein(a); Non-HDL-C, non-high-density lipoprotein cholesterol; POR, proportional odds ratio.

We next asked whether the inverse association was also observed for imaging-defined atherosclerosis in SCAPIS, an independent population-based imaging cohort. A lower genetically proxied NLRP3 activity score was associated with higher coronary artery calcification (*β* = 0.31, 95% CI 0.02 to 0.60, *P* = 0.034) and higher coronary plaque burden, measured as the segment involvement score on CT angiography (proportional odds ratio = 2.55, 95% CI 1.20 to 5.41, *P* = 0.015), with a directionally consistent but weaker association for carotid plaque (proportional odds ratio = 1.88, 95% CI 0.96 to 3.68, *P* = 0.066) (Figure 3B). The higher CAD risk associated with a lower NLRP3 activity score is therefore mirrored in imaging-defined atherosclerotic burden.

### Mediation of the coronary artery disease association by cardiometabolic risk factors

To investigate potential mechanisms underlying the higher CAD risk, we examined associations of the NLRP3 activity score with established cardiometabolic risk factors^49,59^. A lower genetically proxied NLRP3 activity score was associated with several adverse cardiometabolic traits, including higher ApoB levels (*β* = 0.14, 95% CI 0.06 to 0.22, *P* = 0.00078), higher SBP (*β* = 0.11, 95% CI 0.05 to 0.16, *P* = 0.00017), and higher T2D risk (OR = 1.26, 95% CI 1.09 to 1.46, *P* = 0.002), but showed no association with body mass index, smoking, or alcohol consumption (Figure 3C, Supplementary Table 8). To quantify how much of the association with higher CAD risk could be attributed to these cardiometabolic pathways, we performed mediation analysis using ApoB, SBP, and T2D as mediators, with each mediator-to-CAD effect estimated in mutually adjusted MVMR (Figure 3D). Conditional F-statistics were 15.4 for ApoB, 36.7 for SBP, and 37.2 for T2D, indicating strong instruments. Together, the mediators accounted for approximately 87% (95% CI 34%–459%) of the total association between lower genetically proxied NLRP3 activity score and higher CAD risk, with the largest indirect effect attributable to SBP (OR = 1.08, 95% CI 1.04 to 1.13) and smaller contributions from ApoB (OR = 1.05, 95% CI 1.02 to 1.08) and T2D (OR = 1.04, 95% CI 1.02 to 1.07) (Supplementary Table 9). Sequentially accounting for SBP, ApoB, and T2D attenuated the association, leaving a residual association that was no longer statistically significant (OR = 1.03, 95% CI 0.85 to 1.24, *P* = 0.79) (Figure 3E, Supplementary Table 9). Most of the higher CAD risk associated with a lower NLRP3 activity score was therefore attributable to adverse cardiometabolic effects, with little evidence of a residual direct effect.

### Further validation of the NLRP3 activity score and robustness of the coronary artery disease association

We next examined whether the NLRP3 activity score and its association with CAD could be artefacts of the instrument rather than reflect NLRP3 biology. To test whether the inflammatory signal at the *NLRP3* locus is driven by *NLRP3* itself, we examined rare coding variants in the UK Biobank. Carriers of pLOF variants in *NLRP3* had significantly lower CRP (*β* = -0.023, 95% CI -0.032 to -0.015, *P* = 9.4×10^-8^), GlycA (*β* = -0.018, 95% CI -0.027 to -0.010, *P* = 2.8×10^-5^), and neutrophil counts (*β* = -0.012, 95% CI -0.021 to -0.0036, *P* = 5.7×10^-3^) than non-carriers (Figure 4A). Conversely, although carrier numbers were small, the three carriers of the gain-of-function variant had markedly elevated CRP and higher GlycA concentrations and neutrophil counts (Figure 4B), consistent with enhanced NLRP3 inflammasome activity. pLOF burden tests of the other protein-coding genes within ±110 kb of *NLRP3* showed no associations with these biomarkers (Supplementary Figure 3), supporting *NLRP3* as the gene driving the inflammatory biomarker associations at this locus.

**Figure 4.**
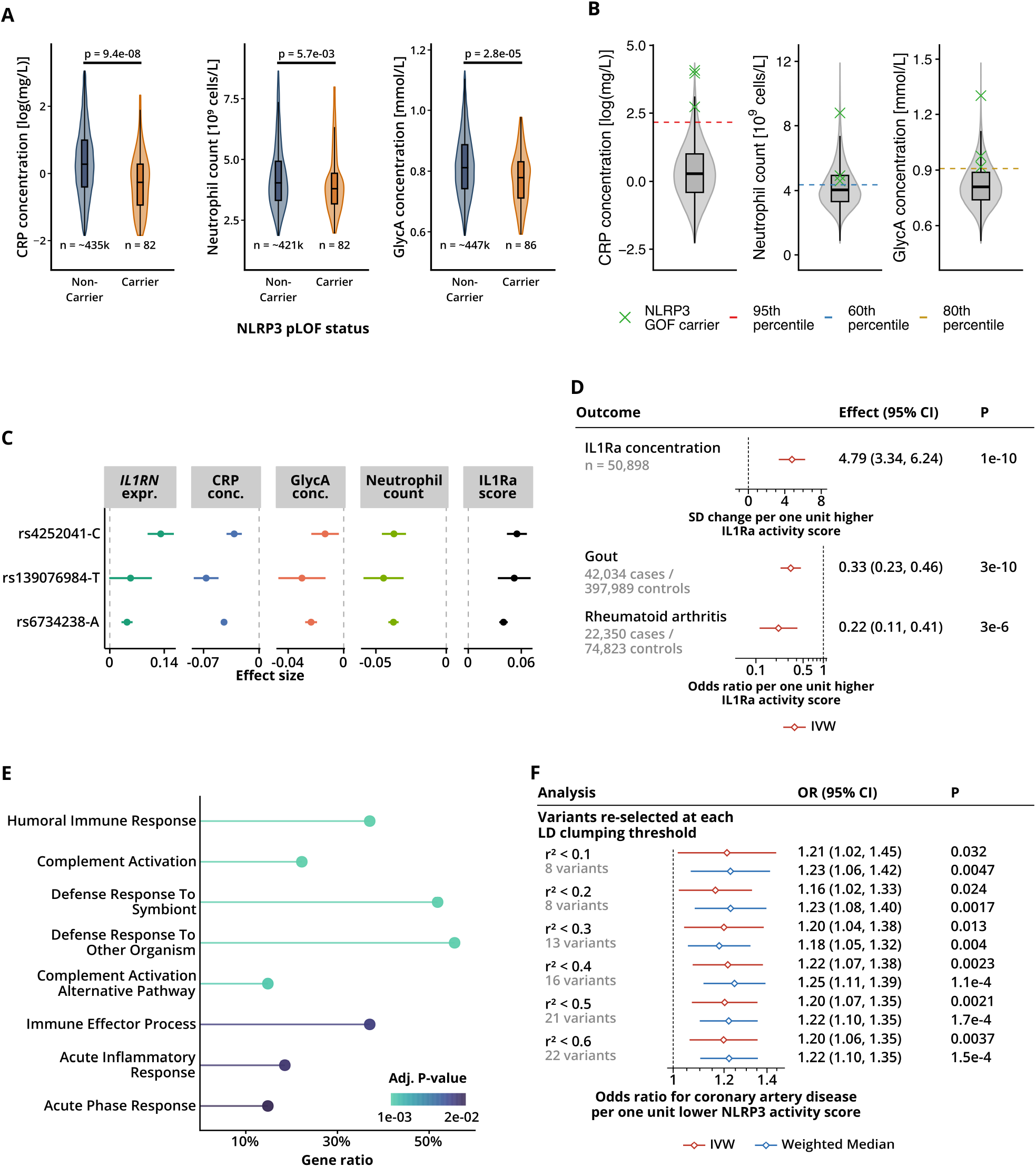
Supporting analyses for the NLRP3 activity score and robustness of the coronary artery disease association. **(A)** Violin plots comparing circulating CRP, neutrophil count, and GlycA levels between carriers and non-carriers of predicted loss-of-function variants in *NLRP3*. The displayed distributions exclude observations outside the 1st and 99th percentiles for visualization. Inset boxplots show the median and interquartile range, with whiskers extending to the most extreme displayed observations within 1.5 times the interquartile range. *P* values are from SAIGE-GENE+ gene-based burden tests. **(B)** Distribution of circulating CRP, neutrophil count, and GlycA levels in UK Biobank participants, with carriers of the *NLRP3* gain-of-function variant highlighted. Dashed lines indicate reference percentiles. **(C)** Per-allele effect sizes and 95% confidence intervals of variants at the *IL1RN* locus on *IL1RN* expression, CRP, GlycA, neutrophil count, and the genetically proxied IL1Ra activity score. The effect allele, shown after each rsID, is the allele that raises *IL1RN* expression. **(D)** Forest plot of MR estimates for the effect of a higher IL1Ra activity score on circulating IL1Ra protein concentration, gout, and rheumatoid arthritis. The protein estimate is reported as SD change per one-unit higher score and disease estimates as odds ratios. Estimates are from random-effects IVW MR only, as only three variants were available, making weighted-median analysis an uninformative sensitivity analysis. **(E)** Over-representation analysis of the 27 circulating proteins decreased with a lower genetically proxied NLRP3 activity score in proteome-wide MR using a one-sided hypergeometric test with all 2,922 tested proteins as background. Gene Ontology Biological Process terms with adjusted *P* < 0.05 are shown. The x-axis shows the gene ratio, calculated as the proportion of significant input proteins annotated to each GO term. Colour indicates the adjusted *P* value. **(F)** Forest plot of MR estimates for the effect of a lower NLRP3 activity score on CAD risk across alternative LD clumping thresholds, with variants re-selected at each threshold. The number of variants at each threshold is shown in the figure. GO, Gene Ontology; GOF, gain-of-function.

As a methodological positive control, we asked whether our activity score MR approach could recover known drug-target biology at the *IL1RN* locus. *IL1RN* encodes IL1Ra, an endogenous inhibitor of interleukin-1 signalling^60^, and recombinant IL1Ra (anakinra) is approved for rheumatoid arthritis (RA) ^60^ and effective in acute gout flares^61^. We therefore constructed an IL1Ra activity score from *IL1RN* expression, CRP, GlycA and neutrophil count (Figure 4C). Because IL1Ra is directly measurable in plasma, we could test the score against the protein whose activity it is meant to proxy. A higher genetically proxied IL1Ra activity score was associated with higher circulating IL1Ra protein concentration (*β* = 4.79, 95% CI 3.34 to 6.24, *P* = 1×10^-10^). Furthermore, consistent with the therapeutic effects of anakinra, a higher IL1Ra activity score was also associated with lower risk of RA (OR = 0.22, 95% CI 0.11 to 0.41, *P* = 3×10^-6^) and gout (OR = 0.33, 95% CI 0.23 to 0.46, *P* = 3×10^-10^) (Figure 4D, Supplementary Table 10).

To characterize the broader inflammatory pathways associated with the NLRP3 activity score, we performed proteome-wide MR. A lower genetically proxied NLRP3 activity score was associated with lower levels of 27 and higher levels of seven circulating proteins after multiple-testing correction (Supplementary Table 11). Over-representation analysis of the 27 proteins with lower levels showed enrichment for acute-phase and complement-activation pathways (Figure 4E), consistent with the hepatic acute-phase response downstream of NLRP3-driven IL-1β and IL-6 signalling. In contrast, the seven proteins with higher levels were enriched only for amino-acid metabolic processes, driven by three proteins (Supplementary Table 12).

Finally, we tested whether the CAD association depended on how the instrument was constructed. The association remained significant across LD clumping thresholds from *r*^2^ = 0.1 to 0.6 (ORs 1.16–1.22, *P* ≤ 0.032) and was directionally consistent in leave-one-out analyses (Figure 4F, Supplementary Figure 4, Supplementary Table 13). A conventional instrument based on *NLRP3* expression alone (12 variants, F-statistic = 65.0) gave a CAD estimate in the same direction (OR = 1.027 per SD lower expression, 95% CI 0.998 to 1.056, P = 0.070), but the alleles associated with lower *NLRP3* expression were not always associated with lower inflammatory biomarkers (Cochran’s Q P ≤ 2×10⁻¹³), and this instrument was not significantly associated with IL-1β or IL-18 (P ≥ 0.11). The CAD estimate was larger when *NLRP3* expression was proxied by the eight variants used in the NLRP3 activity score (OR = 1.044 per SD, 95% CI 1.003 to 1.087), consistent with the CAD association being carried by the variants whose expression effects are accompanied by lower inflammation (Supplementary Information, Supplementary Table 13).

Collectively, these findings identify *NLRP3* as the gene underlying the inflammatory signal at this locus and show that the CAD association is consistent across sensitivity analyses.

### Genetic associations of the NLRP3 activity score across diseases under investigation for NLRP3 inhibition

Beyond CAD, we explored whether the NLRP3 activity score was associated with 15 other diseases for which NLRP3 inhibitors have been, or currently are, under clinical investigation (Supplementary Table 14). A lower genetically proxied NLRP3 activity score was nominally associated with lower risk of gout (OR = 0.64, 95% CI 0.47 to 0.87, *P* = 0.0052), pericarditis (OR = 0.22, 95% CI 0.06 to 0.72, *P* = 0.012), and Parkinson’s disease (PD) (OR = 0.48, 95% CI 0.24 to 0.95, *P* = 0.036) (Figure 5). Type 2 diabetes was the one indication where lower genetically proxied NLRP3 activity was associated with higher risk, and this estimate is reported above as a mediator of the CAD association. No clear associations were observed for the other indications examined (Supplementary Table 15). Together, these exploratory findings provide human genetic evidence consistent with potential benefit of lower NLRP3 activity in selected inflammatory indications.

**Figure 5.**
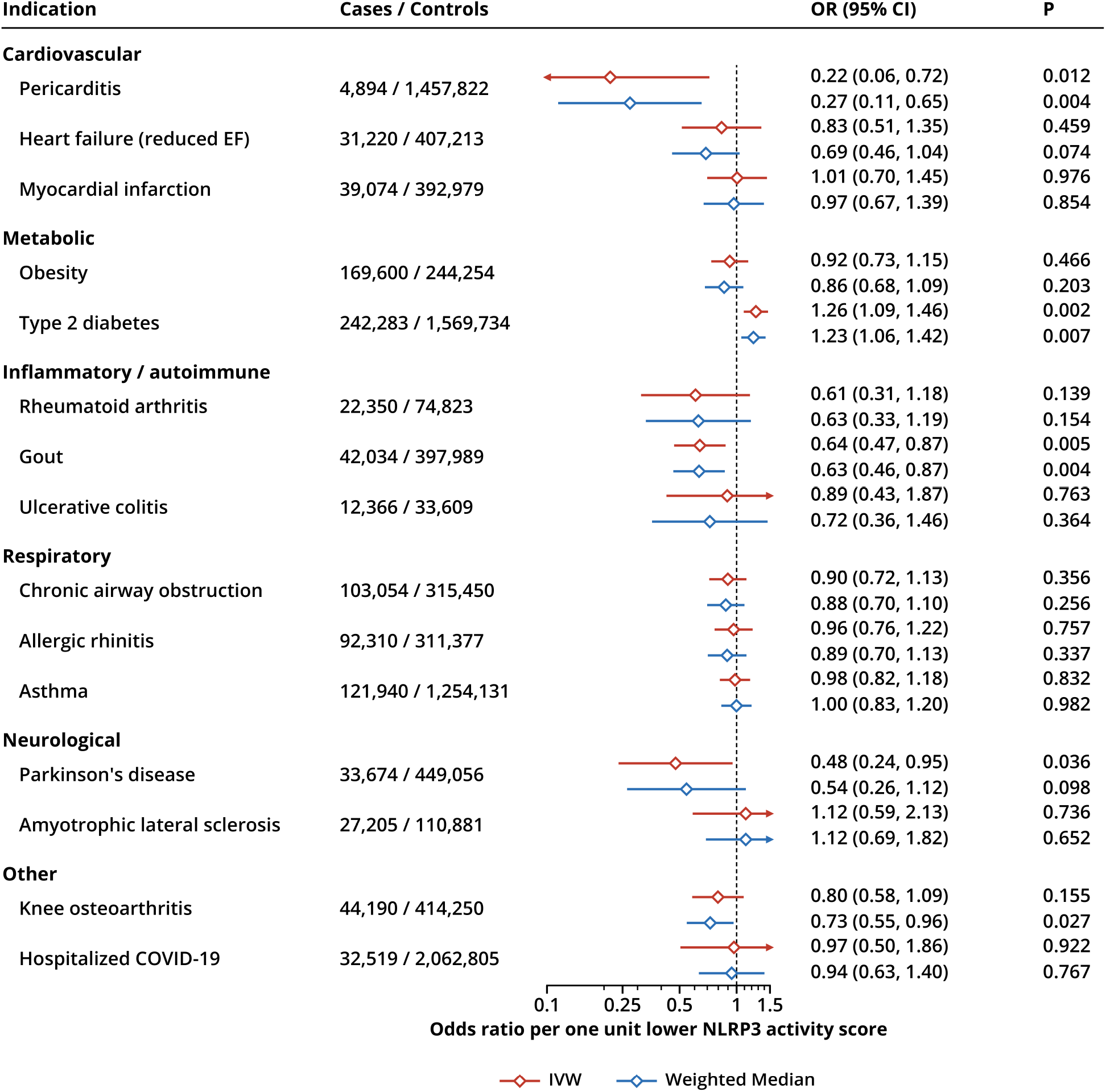
NLRP3 activity score associations across diseases under investigation for NLRP3 inhibition. Forest plot showing MR estimates for the association of a lower NLRP3 activity score with diseases for which NLRP3 inhibitors have been or are being evaluated in clinical trials. Outcomes are grouped by disease area. Estimates are shown as odds ratios per one-unit lower NLRP3 activity score, from inverse-variance weighted and weighted median MR. Parkinson’s disease cases comprise 15,056 diagnosed cases and 18,618 UK Biobank proxy cases. EF, ejection fraction.

## Discussion

In this drug-target MR study, we investigated whether lower genetically proxied NLRP3 activity is associated with CAD risk. Because NLRP3 activity cannot be measured directly in large population datasets^62^, we constructed a composite genetic score combining *NLRP3* expression with downstream inflammatory biomarkers. A lower genetically proxied NLRP3 activity score was associated with lower circulating IL-1β, IL-18, and IL-6, consistent with the expected downstream effects of lower NLRP3 activity. Despite these lower inflammatory markers, a lower NLRP3 activity score was associated with higher CAD risk, and with greater coronary artery calcification and coronary plaque burden. Mediation analyses suggested that this association was largely accounted for by adverse cardiometabolic effects, including higher SBP, higher levels of proatherogenic lipids and ApoB, and greater T2D risk, with little evidence of a residual direct association after accounting for these pathways. In contrast, a lower NLRP3 activity score was nominally associated with lower risk of gout, pericarditis and Parkinson’s disease.

Our findings are surprising in light of the CANTOS trial, where canakinumab reduced recurrent cardiovascular events through inhibition of IL-1β, a downstream effector of NLRP3 inflammasome activation^2,63^. Post hoc analyses of CANTOS showed that canakinumab lowered IL-6 but not IL-18, and that residual levels of both cytokines predicted recurrent events^64^. On this basis, NLRP3 inhibition was proposed to offer greater benefit than IL-1β blockade alone, because it would lower IL-18 in addition to IL-1β, and direct IL-6 inhibition was proposed as an alternative strategy ^64^. This latter strategy did not reduce cardiovascular events in the ZEUS trial among patients with cardiovascular and chronic kidney disease^8^ but is still being tested after acute myocardial infarction (ARTEMIS, NCT06118281). In our analysis, a lower NLRP3 activity score was associated with lower IL-1β, IL-18 and IL-6, yet with higher CAD risk, suggesting that the cardiovascular consequences of lower NLRP3 activity cannot be inferred from its anti-inflammatory effects alone. Experimental evidence is also mixed, as NLRP3 deficiency reduced atherosclerosis in one mouse model^14^, but not in another^65^.

A potential explanation for the higher CAD risk is the adverse cardiometabolic profile associated with lower genetically proxied NLRP3 activity. The ApoB association is consistent with evidence from genetic proxies of IL-1α/β inhibition, which were associated with lower IL-6 and CRP but higher levels of proatherogenic lipids and higher coronary heart disease risk^66^. In RA, IL-6 receptor blockade and TNF inhibition can also raise lipids^67^. The T2D association contrasts with the rationale for testing NLRP3 inhibitors in T2D, but the experimental evidence behind it is mixed. NLRP3 deficiency protected mice against obesity-induced insulin resistance^68^, whereas other studies found no protection^69^ or worse fatty liver disease^70,71^. IL-18 offers one possible explanation, as lower NLRP3 activity was associated with lower IL-18 and its experimental loss impairs insulin sensitivity and energy homeostasis ^72^. The SBP association similarly contrasts with experimental evidence that NLRP3 inhibition lowers blood pressure and renal damage in hypertension^73^. The cardiometabolic effects associated with lifelong lower NLRP3 activity are therefore only partly anticipated by existing experimental and pharmacological evidence. Trials of NLRP3 inhibitors should monitor blood pressure, lipid, and glycaemic effects.

Although our findings do not support lowering NLRP3 activity to reduce CAD risk in the general population, benefit may still exist in selected subgroups with inflammasome-driven cardiovascular risk. Clonal haematopoiesis provides one such setting, with experimental and human data implicating NLRP3-dependent IL-1β signalling in clonal haematopoiesis-associated cardiovascular disease^74^. In an exploratory genomic substudy of CANTOS, participants harbouring *TET2* variants showed a greater reduction in major adverse cardiovascular events when treated with canakinumab^75^.

Our findings differ from those of Schunk *et al*.^18^. Using a single intronic variant (rs10754555) associated with *NLRP3* expression, they reported that higher NLRP3 activity was associated with higher CAD prevalence among people younger than 60 years of age and with higher cardiovascular mortality across primary- and secondary-prevention cohorts. The variant was associated with higher *NLRP3*expression and inflammasome activation in functional experiments, but not with lipid parameters. By contrast, our NLRP3 activity score integrates information across eight variants that tag rs10754555 only weakly, with a maximum *r²* of 0.26 in the INTERVAL reference panel, and was additionally associated with cardiometabolic traits. Because these cardiometabolic associations act on CAD in the opposite direction to the inflammatory associations, they may explain the discordance. The age- and metabolic-context dependence reported by Schunk *et al.* further suggests that the cardiovascular consequences of NLRP3-related genetic variation may depend on the population studied.

Beyond CAD, the associations with gout and pericarditis are consistent with established NLRP3–IL-1 biology. Monosodium urate crystals are canonical NLRP3 activators in gout^76^. In recurrent pericarditis, IL-1 blockade is effective^77^ but relies on expensive injectable biologics. An oral NLRP3 inhibitor has entered phase 2 testing for this indication (NCT06836232), and our estimate lends genetic support to that approach. The PD association contrasts with a recent genetic study that used expression-based instruments for NLRP3 inflammasome-related genes and found no evidence for a role of the NLRP3 inflammasome in PD risk^78^. Our score integrates multiple variants at the *NLRP3* locus with downstream inflammatory readouts, which may contribute to the differing findings. Overall, these exploratory disease associations are consistent with the biological relevance of the NLRP3 activity score beyond CAD but require replication.

We acknowledge that our study has limitations. Although *cis*-restriction reduces the likelihood of horizontal pleiotropy, it cannot eliminate it, because variants may influence several biological processes through mechanisms not fully captured by the intended exposure^79^. The cardiometabolic associations could therefore reflect horizontal rather than vertical pleiotropy, a possibility consistent with the non-zero MR-Egger intercept for CAD at the primary clumping threshold (*r*² = 0.1). However, if the variants affected SBP, ApoB, or T2D through pathways other than NLRP3 activity, their effects on these traits would generally not be proportional to their effects on the NLRP3 activity score. MR-Egger regression showed no such departure from proportionality for any of the three traits, or for CAD at relaxed clumping thresholds (Egger intercept *P* > 0.12). In addition, rare loss-of-function and gain-of-function variants in *NLRP3* affected the inflammatory biomarkers in the expected directions, and no neighbouring gene showed comparable associations, supporting *NLRP3* as the causal gene at this locus. Bias from participant overlap is also possible, as several exposure and outcome datasets include UK Biobank participants, although with strong instruments (F = 92) the bias is expected to be minimal^80^. Another limitation is that genetic variants capture small, lifelong differences in NLRP3 activity, whereas pharmacological inhibition produces a larger perturbation over a defined treatment period. The adverse cardiometabolic effects observed here may not fully develop within that period, so trials could find less excess CAD risk than our genetic estimates suggest^81^. Our estimates also apply to a general population, whereas clinical studies of direct NLRP3 inhibition in CAD have focused on patients with established coronary artery disease, in whom the cardiometabolic pathways implicated here are aggressively treated with lipid-lowering and antihypertensive therapy. Statistical power is a further consideration. The residual direct effect on CAD, after accounting for SBP, ApoB, and T2D, was close to the null, but its confidence interval remains compatible with a modest effect in either direction. Relatedly, for several disease outcomes, the available case numbers may have been insufficient to detect modest causal effects, and null findings should therefore be interpreted with caution. Our instrument is also derived from inflammatory biomarkers measured in peripheral blood and may not reflect NLRP3 activity in disease-relevant tissues such as the coronary arteries or the brain. Establishing these tissue-specific effects will require complementary evidence, including tissue-specific molecular data and pharmacodynamic biomarker measurements in clinical studies. Finally, the NLRP3 activity score and all disease outcome analyses were based on European-ancestry data, which limits generalizability.

In conclusion, a lower genetically proxied NLRP3 activity score was associated with higher CAD risk, with mediation analyses suggesting that this association was largely accounted for by adverse cardiometabolic effects. These findings reflect lifelong genetic variation in the general population and do not preclude benefit from pharmacological inhibition in selected patients with inflammasome-driven cardiovascular risk. They provide no genetic support for lowering NLRP3 activity to reduce CAD risk and argue for monitoring cardiometabolic effects in trials of NLRP3 inhibitors.

## Data Availability

All GWAS summary statistics analysed in this study are publicly available from the sources listed in Supplementary Tables 1-4. Individual-level data from UK Biobank and the other contributing studies are available to approved researchers through their respective access procedures. This study used data from the All of Us Research Program's Controlled Tier Dataset v8, available to authorized users on the Researcher Workbench. The code used for all analyses and figures is available at https://github.com/nickhir/NLRP3-drug-target-MR.

## Acknowledgements

This research has been conducted using the UK Biobank Resource under Application Number 608471. We gratefully acknowledge All of Us participants for their contributions, without whom this research would not have been possible. We also thank the National Institutes of Health’s All of Us Research Program for making available the participant data examined in this study. We want to acknowledge the participants and investigators of the FinnGen study. We thank the participants and investigators of UK Biobank, INTERVAL, the Million Veteran Program, SCAPIS and deCODE genetics, and all consortia that made their summary statistics publicly available. This work was performed using resources provided by the Cambridge Service for Data Driven Discovery (CSD3) operated by the University of Cambridge Research Computing Service (www.csd3.cam.ac.uk), provided by Dell EMC and Intel using Tier-2 funding from the Engineering and Physical Sciences Research Council (capital grant EP/T022159/1), and DiRAC funding from the Science and Technology Facilities Council (www.dirac.ac.uk). The authors used Claude (Anthropic) to assist with writing, editing, and code generation. All AI-assisted code and text were critically reviewed and edited by the authors, who take full responsibility for the integrity and accuracy of the analyses and the manuscript content.

## Data Availability

All GWAS summary statistics analysed in this study are publicly available from the sources listed in Supplementary Tables 1–4. Individual-level data from UK Biobank and the other contributing studies are available to approved researchers through their respective access procedures. This study used data from the All of Us Research Program’s Controlled Tier Dataset v8, available to authorized users on the Researcher Workbench. The code used for all analyses and figures is available at https://github.com/nickhir/NLRP3-drug-target-MR.

## Ethical Approval

This study used summary statistics from published genome-wide association studies, each of which obtained ethical approval and participants’ informed consent as described in the original publications; no further approval was required for their use. Individual-level UK Biobank data were analysed under Application Number 608471. UK Biobank has Research Tissue Bank approval from the North West Multi-centre Research Ethics Committee (11/NW/0382, renewed as 16/NW/0274 and 21/NW/0157), and all participants gave written informed consent. The INTERVAL study was approved by the National Research Ethics Service (11/EE/0538), and all participants gave informed consent. All of Us participants provided informed consent. Only All of Us summary statistics were used, accessed through the All of Us Researcher Workbench. The study was conducted in accordance with the Declaration of Helsinki.

## Pre-registered Clinical Trial Number

Not applicable.

## Supplementary Data

**Supplementary Figure 1.**
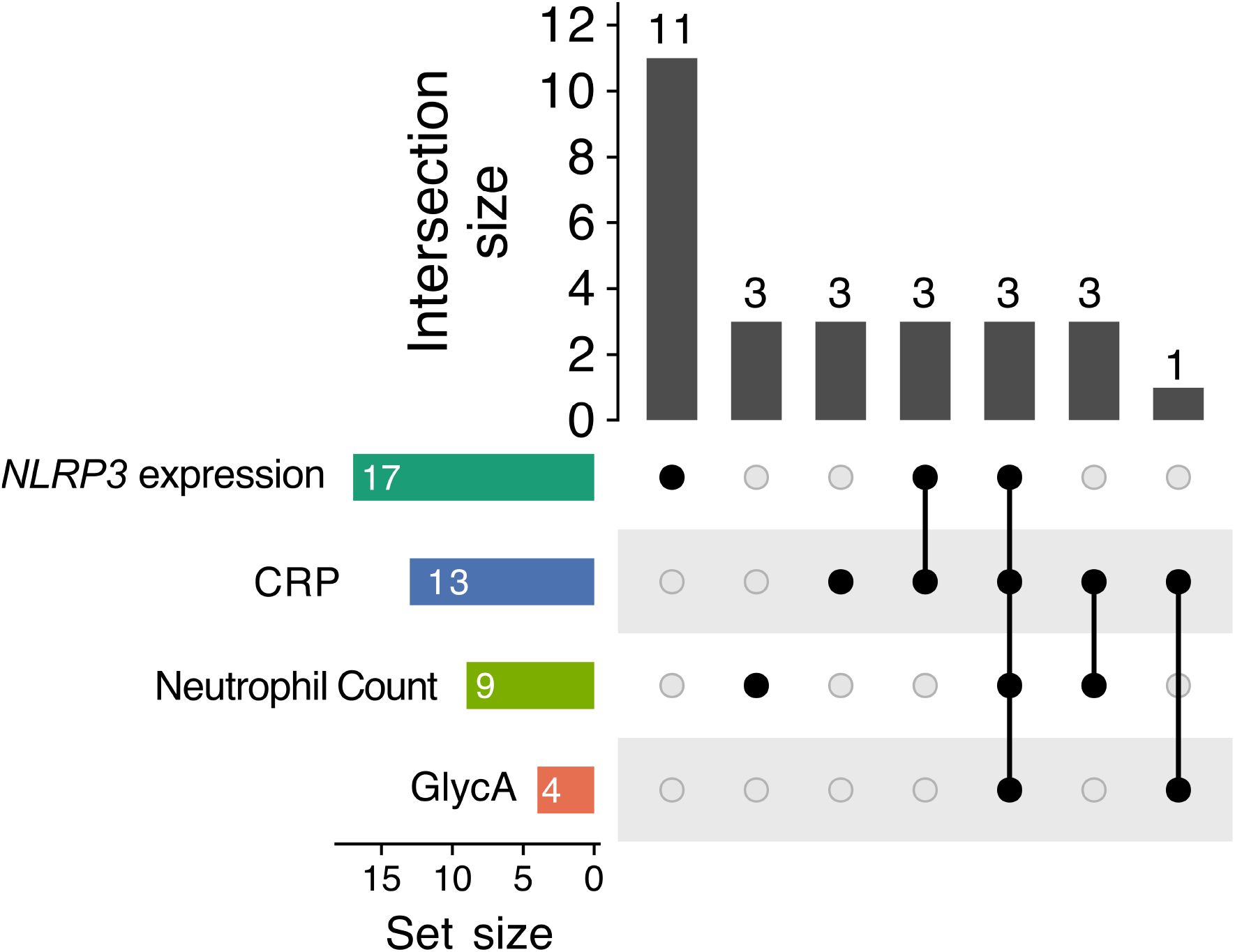
Overlap of variants associated with the four traits used to construct the NLRP3 activity score. UpSet plot showing the overlap of variants within ±150 kb of *NLRP3* associated at genome-wide significance with whole-blood *NLRP3* expression, circulating CRP and GlycA levels, or blood neutrophil count after LD clumping at r²<0.1. Set size indicates the number of clumped genome-wide significant variants per trait, and intersection size indicates the number of variants belonging to each specific combination of traits. Signals associated with at least two traits and with effect estimates available for all four were retained, yielding the eight variants used to construct the NLRP3 activity score. CRP, C-reactive protein; GlycA, glycoprotein acetylation; LD, linkage disequilibrium.

**Supplementary Figure 2.**
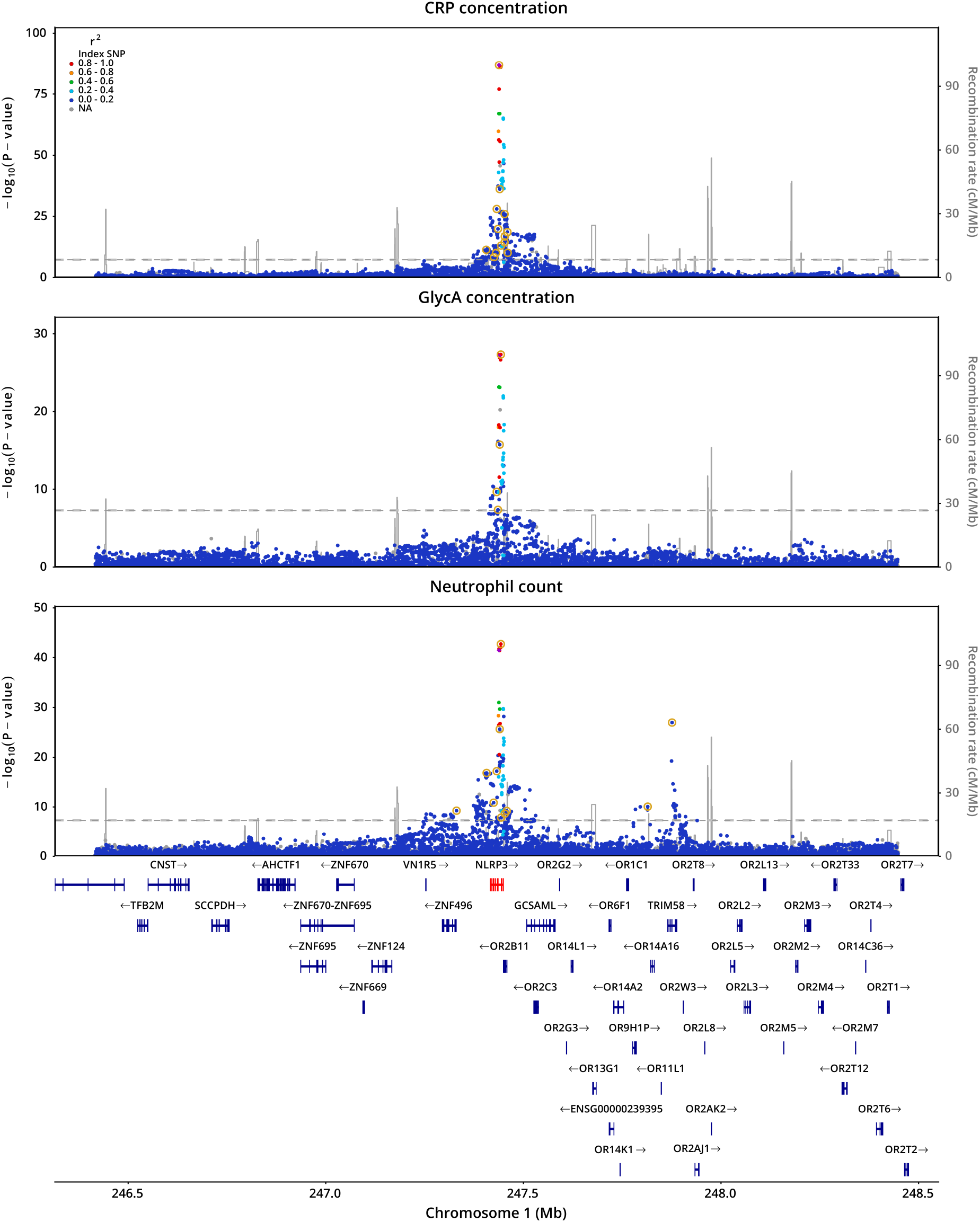
Regional association plots for inflammatory traits within ±1 Mb of the *NLRP3* locus. Regional association plots for CRP concentration, GlycA concentration, and neutrophil count within a ±1 Mb window surrounding *NLRP3*. Variants highlighted with gold circles are approximately independent genome-wide significant variants identified after LD clumping at *r*² < 0.1. Within this region, the shared association signal across traits was centred on the *NLRP3* locus. Additional neutrophil count association signals were not accompanied by corresponding associations with CRP or GlycA. CRP, C-reactive protein; GlycA, glycoprotein acetylation; LD, linkage disequilibrium.

**Supplementary Figure 3.**
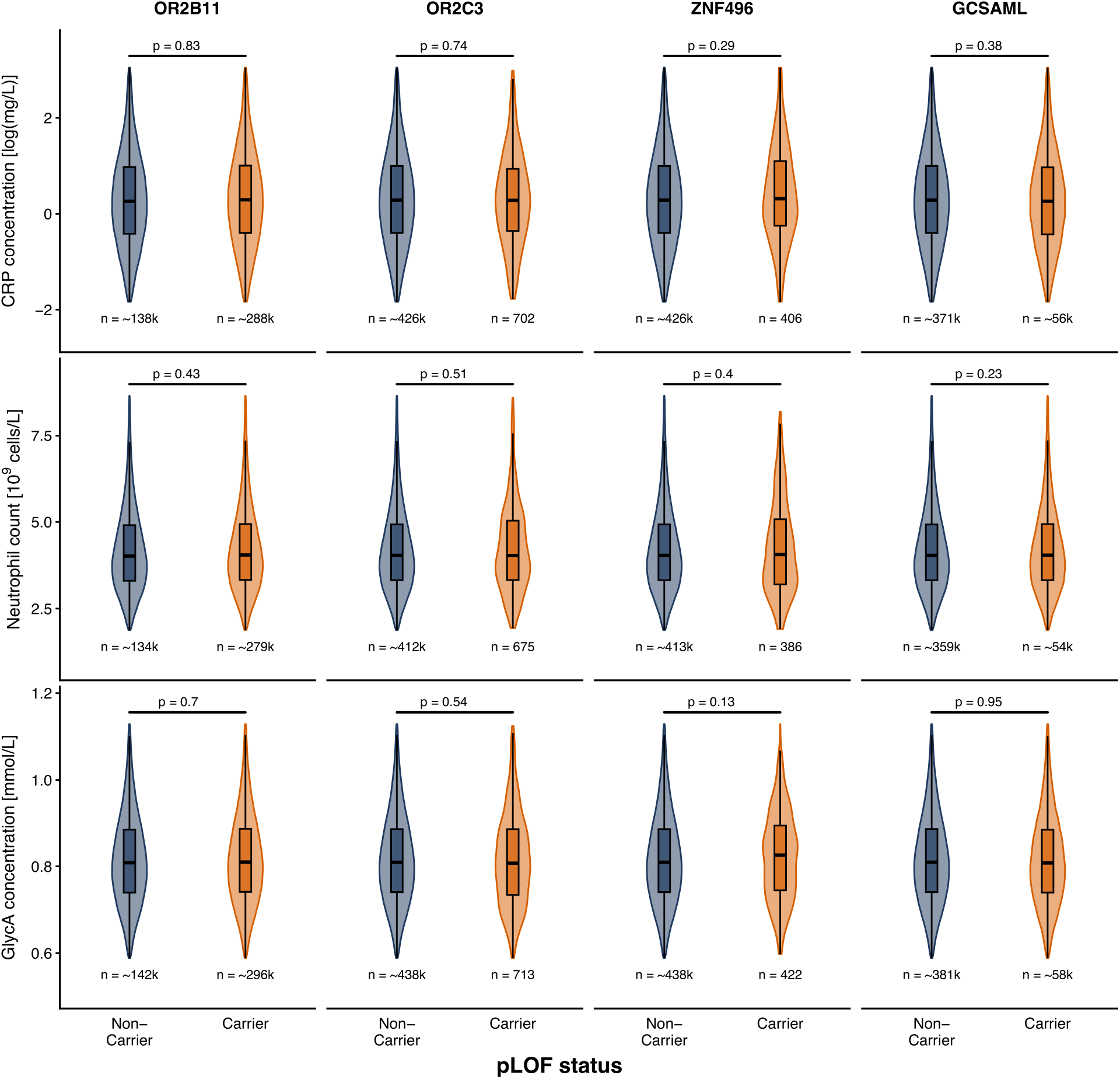
pLOF burden analysis of protein-coding genes surrounding *NLRP3*. Violin plots comparing circulating CRP, neutrophil count, and GlycA levels between carriers and non-carriers of predicted loss-of-function (pLOF) variants in all protein-coding genes within ±110 kb of *NLRP3*. The displayed distributions exclude observations outside the 1st and 99th percentiles for visualization. Inset boxplots show the median and interquartile range, with whiskers extending to the most extreme displayed observations within 1.5 times the interquartile range. P-values are from SAIGE-GENE+ gene-based burden tests. CRP, C-reactive protein; GlycA, glycoprotein acetylation; pLOF, predicted loss-of-function.

**Supplementary Figure 4.**
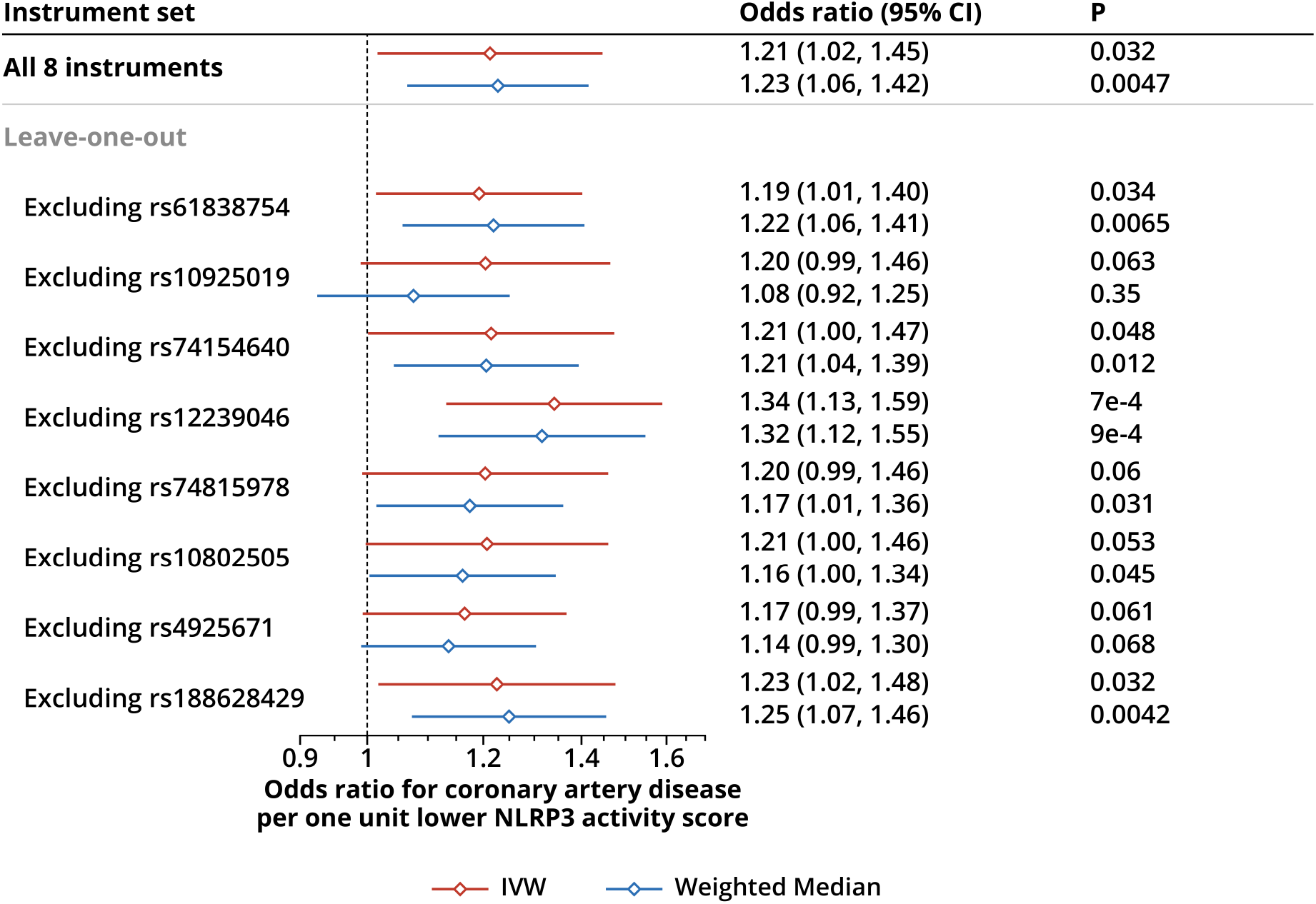
Sensitivity analyses for the association of the NLRP3 activity score with coronary artery disease risk. Forest plot showing MR estimates for the effect of a one-unit lower NLRP3 activity score on CAD risk using the full eight-variant instrument set and sequential leave-one-out analyses excluding each variant in turn. Estimates for the full and leave-one-out instrument sets are shown using inverse-variance weighted and weighted median MR. Odds ratios and 95% confidence intervals are based on the fixed-effect meta-analysis of the four contributing CAD GWAS. CAD, coronary artery disease; CI, confidence interval; GWAS, genome-wide association study; IVW, inverse-variance weighted; MR, Mendelian randomization.

## Supplementary Information

### Choice of traits for the NLRP3 activity score

No assay of NLRP3 inflammasome activity is available at population scale^62^. We therefore combined variant effects on *NLRP3* expression, which anchors the score to the target gene, with their effects on downstream inflammatory traits. We did not use expression alone because inflammasome assembly depends on post-translational signals in addition to transcriptional priming, so *NLRP3* mRNA levels may not reflect inflammasome output^9^. Consistent with this, alleles that lower *NLRP3* expression differ in whether they lower downstream inflammatory traits (see below).

The most proximal circulating readouts of NLRP3 inflammasome activation are IL-1β and IL-18. Further downstream are IL-6 and the acute-phase response, reflected in CRP, GlycA and fibrinogen, together with increased granulopoiesis, reflected in neutrophil count^9–12^. GlycA, a composite NMR signal from several glycosylated acute-phase proteins including α1-acid glycoprotein, haptoglobin and α1-antitrypsin^11^, extends the acute-phase readout beyond CRP. Of these readouts, CRP, GlycA and neutrophil count have by far the largest GWAS (more than 430,000 participants each^19–21)^, and each shows genome-wide significant associations at the *NLRP3* locus. We therefore used them to construct the score. While IL-1β and IL-18 more directly reflect inflammasome activation, they and IL-6 were measured in only about 50,000 participants in the UK Biobank Pharma Proteomics Project^23^, giving far less power to detect the small effects of individual *NLRP3* variants. Holding them out of construction instead allowed them to serve as an independent test of whether the score captures inflammasome output.

CRP, GlycA and neutrophil count are not specific to the NLRP3 inflammasome. Specificity was instead addressed through the design: instruments were restricted to *cis* variants at *NLRP3*, variants associated with the expression of other genes were excluded, and we tested whether these traits shared a causal variant with *NLRP3* expression and were shifted by rare loss- and gain-of-function variants in *NLRP3* but not by loss-of-function variants in neighbouring genes.

### Construction of the NLRP3 and IL1Ra activity scores

To construct the NLRP3 activity score, we used genetic associations for four traits related to NLRP3 inflammasome activation: whole-blood *NLRP3* expression, circulating CRP and GlycA levels, and blood neutrophil count. For each trait, variants within *±*150 kb of *NLRP3* were identified and LD-clumped using PLINK2^82^ (version 2.00a6) with a 250 kb clumping window, a genome-wide significance threshold of *P*∈5 *×*10^−8^, and a primary clumping threshold of *r*^2^ ∈0.1. The LD reference panel comprised 11,863 European-ancestry whole-genome sequences from INTERVAL. Only single-nucleotide variants were used, and variants whose effect-allele frequency in the panel differed from the neutrophil-count GWAS frequency by more than 0.05 were excluded from the reference panel. Because standard clumping may select different lead variants for different traits even when these tag the same underlying signal, we next identified variants in high LD with each trait-specific lead variant using *r*^2^ >0.95 within a 40 kb window. These high-LD variant sets were treated as signal blocks, and blocks were merged across traits if they shared at least one variant. Merged blocks represented in at least two of the four traits were retained for score construction, of which there were ten. For each retained block, we selected a single representative variant, prioritizing variants available across all four datasets and then choosing the variant with the lowest CRP association *P*-value. Where the representative variant was absent from one or more datasets, we substituted a proxy within 50 kb, provided *r*² was at least 0.9. Two representatives were still missing from at least one dataset after this substitution and were excluded, leaving eight variants for construction of the NLRP3 activity score (Supplementary Figure 1). We further required instruments to be specific to the target gene. Using INTERVAL whole-blood expression quantitative trait locus (eQTL) summary statistics, a variant was excluded if it was associated (P < 1×10⁻³) with expression of another gene tested in *cis*, and this association persisted (P < 1×10⁻³) after conditioning on that gene’s lead eQTL. This increased the specificity of the instruments for their target gene.

For each retained variant *j*, trait-specific genetic effect estimates were assembled into the vector 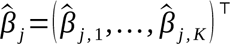, where *K*=4 denotes the four traits used to construct the score. We centred and scaled the trait-specific effect estimates to have mean zero and unit variance across variants before applying PCA to the resulting variant-by-trait matrix. Loadings from the first principal component were extracted and used as fixed weights w=( w_1_ ,…, w_k_ )^⊤^ to combine trait-specific variant effects into a single latent factor:

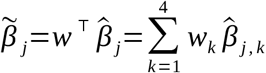

This latent factor was defined as the NLRP3 activity score. Because its raw scale is not directly interpretable, we scaled it to CRP. Specifically, we estimated a scaling constant *c* from a weighted linear regression of the CRP effect estimate *^βj* on the unscaled score effect ∼*βj*, with the intercept constrained to zero and inverse-variance weights based on the CRP standard errors. The scaled variant effect on the NLRP3 activity score was then defined as:

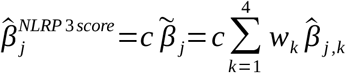

The score was oriented so that lower values corresponded to lower CRP. By construction, a one-unit decrease in the scaled score corresponds approximately to a one standard deviation decrease in CRP.

Standard errors for 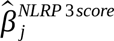were estimated using the delta method^43^. For each variant, we defined a variance-covariance matrix for the trait-specific effect estimates. Because the CRP, GlycA, and neutrophil count GWAS may include overlapping participants, we used a conservative approach and assumed perfectly correlated sampling errors across these biomarker traits. This assumption inflates standard errors and therefore yields conservative inference. As *NLRP3* expression was measured in a distinct cohort, expression quantitative trait loci effect estimates were assumed to be independent of the biomarker effect estimates.

The same procedure was applied at the *IL1RN* locus to construct a genetically proxied interleukin-1 receptor antagonist (IL1Ra) activity score, using *IL1RN* expression and the same blood-based inflammatory traits.

The expression-only instrument was constructed with the same selection pipeline, using INTERVAL whole-blood *NLRP3* expression as the only readout. Of its 12 variants, four are also NLRP3 activity score instruments and a further two are in LD with NLRP3 activity score signals (r² > 0.9). The expression-lowering alleles at rs72771965 and rs4925645 were associated with higher rather than lower CRP and neutrophil count. The instrument therefore averages over variants with opposing downstream effects, which is reflected in highly significant between-variant heterogeneity for all three inflammatory biomarkers (Cochran’s Q P ≤ 2×10⁻¹³). We also estimated the CAD association using the effects of the eight NLRP3 activity score variants on *NLRP3* expression alone.

### Mediation analysis

ApoB was selected as the lipid mediator because it reflects the concentration of atherogenic ApoB-containing lipoprotein particles and provides a parsimonious measure of lipid-related cardiovascular risk, avoiding joint modelling of highly correlated lipid fractions^50^. SBP was preferred to diastolic blood pressure, with which it is highly correlated, because it showed stronger evidence of association with the NLRP3 activity score. The MVMR instruments were selected independently of the *cis*-acting NLRP3 variants used to instrument the activity score. For MVMR, variants were clumped within each trait and then again across traits (r² < 0.001, 10 Mb window, INTERVAL reference panel), so that the combined instrument set of 1,265 variants is approximately independent. Multiplicative random effects were applied to account for heterogeneity across the mediator instruments. Because the mediator GWAS draw on partly overlapping samples, the covariance between their variant effect estimates was estimated from z-scores at variants that were null for both traits (P > 0.05), using one variant from each 100 kb window across the genome so that the variants were approximately independent^83^.

### *NLRP3* rare variant phenotype processing

For *NLRP3* pLOF burden analyses and gain-of-function carrier analyses, CRP levels and GlycA concentrations were adjusted for age, age^2^, sex, age × sex, age^2^ × sex, assessment centre, genotyping array, and the first 20 genetic principal components before inverse-rank normalization. GlycA concentrations were additionally adjusted for technical variation using the ukbnmr package (version 3.3.1)^84^. Blood neutrophil count was adjusted following the blood-cell trait processing framework described by Astle *et al.*, including adjustment for technical variation, age- and sex-related effects, height and weight, smoking, alcohol-related covariates, assessment centre, and genetic principal components before inverse-rank normalization^85^.

### Proteome-wide multiple-testing correction and over-representation analysis

To account for correlation between protein measurements, we estimated the effective number of independent protein tests. To do this, protein measurements were first adjusted for age, age², sex, age × sex, age² × sex, assessment centre, genotyping batch, Olink batch, and the first 20 genetic principal components. Pairwise Pearson correlations were then calculated across adjusted protein measurements, and eigenvalue decomposition of the resulting correlation matrix was used to estimate the effective number of independent tests. The effective number of tests was defined as the number of principal components required to explain 95% of the total variance^58^. This yielded 1,821 effective tests, corresponding to a significance threshold of *P* < 2.7×10^-5^.

To assess whether proteins altered by lower genetically proxied NLRP3 activity were enriched for specific biological processes, we performed over-representation analysis using clusterProfiler (version 4.10.1)^86^. Proteins passing the multiple-testing threshold were tested for enrichment of Gene Ontology Biological Process terms obtained using msigdbr (version 25.1.1)^87,88^, separately by direction of association. The background set comprised all 2,922 tested proteins. P-values were adjusted using the Benjamini-Hochberg method^89^, as Gene Ontology terms overlap heavily and Bonferroni correction would be overly conservative.

### Selection of disease indications from clinical trial registries

To identify disease indications for which selective NLRP3 inhibitors have entered clinical evaluation, we searched ClinicalTrials.gov, the EU Clinical Trials Register (EudraCT), the EU Clinical Trials Information System (CTIS) and ISRCTN on 28 April 2026 using two complementary strategies.

Search 1 (target-level search): Each registry was searched for the term “NLRP3”, restricted to interventional studies.

Search 2 (drug-name search): Each registry was searched for the following compound names: dapansutrile, OLT1177, DFV890, IFM-2427, selnoflast, RO7486967, RG6418, inzomelid, IZD174, somalix, IZD334, emlenoflast, NT-0796, ruvonoflast, NT-0249, VTX2735, VTX3232, VENT-02, ZYIL1, usnoflast, ACI-19764, AZD4144, HL-400, JTE-162, JT002, GDC-2394, and MCC950.

Results from both searches were merged and de-duplicated, with trials registered in more than one registry counted once. Each record was screened and a trial was included only if the intervention was a compound designed specifically to inhibit NLRP3. In addition, rheumatoid arthritis was included because a selective NLRP3 inhibitor (MCC950/CP-456773) was evaluated in a phase 2 trial but was not developed further after it raised serum liver enzyme levels^90^. The complete list of eligible trials and the disease indications extracted from them are provided in Supplementary Table 14.

## References

1. Ridker PM. Clinician’s Guide to Reducing Inflammation to Reduce Atherothrombotic Risk: JACC Review Topic of the Week. J Am Coll Cardiol 2018;72:3320–3331.

2. Ridker PM, Everett BM, Thuren T, MacFadyen JG, Chang WH, Ballantyne C, et al. Antiinflammatory Therapy with Canakinumab for Atherosclerotic Disease. N Engl J Med 2017;377:1119–1131.

3. Tardif J-C, Kouz S, Waters DD, Bertrand OF, Diaz R, Maggioni AP, et al. Efficacy and Safety of Low-Dose Colchicine after Myocardial Infarction. N Engl J Med 2019;381:2497–2505.

4. Nidorf SM, Fiolet ATL, Mosterd A, Eikelboom JW, Schut A, Opstal TSJ, et al. Colchicine in Patients with Chronic Coronary Disease. N Engl J Med 2020;383:1838–1847.

5. Jolly SS, d’Entremont M-A, Lee SF, Mian R, Tyrwhitt J, Kedev S, et al. Colchicine in Acute Myocardial Infarction. N Engl J Med 2025;392:633–642.

6. Kelly P, Lemmens R, Weimar C, Walsh C, Purroy F, Barber M, et al. Long-term colchicine for the prevention of vascular recurrent events in non-cardioembolic stroke (CONVINCE): a randomised controlled trial. The Lancet 2024;404:125–133.

7. Li J, Meng X, Shi F-D, Jing J, Gu H-Q, Jin A, et al. Colchicine in patients with acute ischaemic stroke or transient ischaemic attack (CHANCE-3): multicentre, double blind, randomised, placebo controlled trial. BMJ 2024;385:e079061.

8. Ridker PM, Lincoff AM, Marx N, Navar AM, Sattar N, Tuttle KR, et al. Interleukin-6 Inhibition with Ziltivekimab for Atherosclerotic Disease. N Engl J Med 2026.

9. Swanson KV, Deng M, Ting JP-Y. The NLRP3 inflammasome: molecular activation and regulation to therapeutics. Nat Rev Immunol 2019;19:477–489.

10. Lee J-K, Kim S-H, Lewis EC, Azam T, Reznikov LL, Dinarello CA. Differences in signaling pathways by IL-1β and IL-18. Proc Natl Acad Sci U S A 2004;101:8815–8820.

11. Bartlett DB, Connelly MA, AbouAssi H, Bateman LA, Tune KN, Huebner JL, et al. A novel inflammatory biomarker, GlycA, associates with disease activity in rheumatoid arthritis and cardio-metabolic risk in BMI-matched controls. Arthritis Res Ther 2016;18:86.

12. Sreejit G, Nooti SK, Jaggers RM, Athmanathan B, Park KH, Al-Sharea A, et al. Retention of the NLRP3 Inflammasome-Primed Neutrophils in the Bone Marrow Is Essential for Myocardial Infarction-Induced Granulopoiesis. Circulation 2022;145:31–44.

13. van der Heijden T, Kritikou E, Venema W, van Duijn J, van Santbrink PJ, Slütter B, et al. NLRP3 Inflammasome Inhibition by MCC950 Reduces Atherosclerotic Lesion Development in Apolipoprotein E–Deficient Mice—Brief Report. Arterioscler Thromb Vasc Biol 2017;37:1457–1461.

14. Duewell P, Kono H, Rayner KJ, Sirois CM, Vladimer G, Bauernfeind FG, et al. NLRP3 inflammasomes are required for atherogenesis and activated by cholesterol crystals. Nature 2010;464:1357–1361.

15. Jiang Y, Qiang Z, Liu Y, Zhu L, Xiao L, Du Z, et al. Diverse functions of NLRP3 inflammasome in PANoptosis and diseases. Cell Death Discov 2025;11:389.

16. Ference BA, Robinson JG, Brook RD, Catapano AL, Chapman MJ, Neff DR, et al. Variation in PCSK9 and HMGCR and Risk of Cardiovascular Disease and Diabetes. N Engl J Med 2016;375:2144–2153.

17. Gregson JM, Freitag DF, Surendran P, Stitziel NO, Chowdhury R, Burgess S, et al. Genetic invalidation of Lp-PLA_2_ as a therapeutic target: Large-scale study of five functional Lp-PLA_2_-lowering alleles. Eur J Prev Cardiol 2017;24:492–504.

18. Schunk SJ, Kleber ME, März W, Pang S, Zewinger S, Triem S, et al. Genetically determined NLRP3 inflammasome activation associates with systemic inflammation and cardiovascular mortality. Eur Heart J 2021;42:1742–1756.

19. Said S, Pazoki R, Karhunen V, Võsa U, Ligthart S, Bodinier B, et al. Genetic analysis of over half a million people characterises C-reactive protein loci. Nat Commun 2022;13:2198.

20. Zoodsma M, Beuchel C, Yasmeen S, Kohleick L, Nepal A, Koprulu M, et al. A genetic map of human metabolism across the allele frequency spectrum. Nat Genet 2025;57:2445–2455.

21. Chen M-H, Raffield LM, Mousas A, Sakaue S, Huffman JE, Moscati A, et al. Trans-ethnic and Ancestry-Specific Blood-Cell Genetics in 746,667 Individuals from 5 Global Populations. Cell 2020;182:1198–1213.e14.

22. Tokolyi A, Persyn E, Nath AP, Burnham KL, Marten J, Vanderstichele T, et al. The contribution of genetic determinants of blood gene expression and splicing to molecular phenotypes and health outcomes. Nat Genet 2025;57:616–625.

23. Sun BB, Chiou J, Traylor M, Benner C, Hsu Y-H, Richardson TG, et al. Plasma proteomic associations with genetics and health in the UK Biobank. Nature 2023;622:329–338.

24. Aragam KG, Jiang T, Goel A, Kanoni S, Wolford BN, Atri DS, et al. Discovery and systematic characterization of risk variants and genes for coronary artery disease in over a million participants. Nat Genet 2022;54:1803–1815.

25. Verma A, Huffman JE, Rodriguez A, Conery M, Liu M, Ho Y-L, et al. Diversity and scale: Genetic architecture of 2068 traits in the VA Million Veteran Program. Science 2024;385:eadj1182.

26. Lu W, Carroll RJ, Solomonson M, Guez J, He MK, Marten DJ, et al. Systematic common and rare variant association testing in 392,030 whole genomes in All of Us. medRxiv 2026. Available from: 10.64898/2026.05.08.26350964.

27. Kurki MI, Karjalainen J, Palta P, Sipilä TP, Kristiansson K, Donner KM, et al. FinnGen provides genetic insights from a well-phenotyped isolated population. Nature 2023;613:508–518.

28. Gummesson A, Lundmark P, Chen QS, Björnson E, Dekkers KF, Hammar U, et al. A genome-wide association study of imaging-defined atherosclerosis. Nat Commun 2025;16:2266.

29. van Rheenen W, van der Spek RAA, Bakker MK, van Vugt JJFA, Hop PJ, Zwamborn RAJ, et al. Common and rare variant association analyses in amyotrophic lateral sclerosis identify 15 risk loci with distinct genetic architectures and neuron-specific biology. Nat Genet 2021;53:1636–1648.

30. Zhou W, Kanai M, Wu K-HH, Rasheed H, Tsuo K, Hirbo JB, et al. Global Biobank Meta-analysis Initiative: Powering genetic discovery across human disease. Cell Genomics 2022;2:100192.

31. Kanai M, Andrews SJ, Cordioli M, Stevens C, Neale BM, Daly M, et al. A second update on mapping the human genetic architecture of COVID-19. Nature 2023;621:E7–E26.

32. Carss K, Halldorsson BV, Hou L, Liu J, Wheeler E, Lo Y, et al. Whole-genome sequencing of 490,640 UK Biobank participants. Nature 2025;645:692–701.

33. Nalls MA, Blauwendraat C, Vallerga CL, Heilbron K, Bandres-Ciga S, Chang D, et al. Identification of novel risk loci, causal insights, and heritable risk for Parkinson’s disease: a meta-analysis of genome-wide association studies. Lancet Neurol 2019;18:1091–1102.

34. Thorolfsdottir RB, Jonsdottir AB, Sveinbjornsson G, Aegisdottir HM, Oddsson A, Stefansson OA, et al. Variants at the Interleukin 1 Gene Locus and Pericarditis. JAMA Cardiol 2024;9:165–172.

35. Ishigaki K, Sakaue S, Terao C, Luo Y, Sonehara K, Yamaguchi K, et al. Multi-ancestry genome-wide association analyses identify novel genetic mechanisms in rheumatoid arthritis. Nat Genet 2022;54:1640–1651.

36. Suzuki K, Hatzikotoulas K, Southam L, Taylor HJ, Yin X, Lorenz KM, et al. Genetic drivers of heterogeneity in type 2 diabetes pathophysiology. Nature 2024;627:347–357.

37. de Lange KM, Moutsianas L, Lee JC, Lamb CA, Luo Y, Kennedy NA, et al. Genome-wide association study implicates immune activation of multiple integrin genes in inflammatory bowel disease. Nat Genet 2017;49:256–261.

38. Graham SE, Clarke SL, Wu K-HH, Kanoni S, Zajac GJM, Ramdas S, et al. The power of genetic diversity in genome-wide association studies of lipids. Nature 2021;600:675–679.

39. Loya H, Kalantzis G, Cooper F, Palamara PF. A scalable variational inference approach for increased mixed-model association power. Nat Genet 2025;57:461–468.

40. Keaton JM, Kamali Z, Xie T, Vaez A, Williams A, Goleva SB, et al. Genome-wide analysis in over 1 million individuals of European ancestry yields improved polygenic risk scores for blood pressure traits. Nat Genet 2024;56:778–791.

41. Pulit SL, Stoneman C, Morris AP, Wood AR, Glastonbury CA, Tyrrell J, et al. Meta-analysis of genome-wide association studies for body fat distribution in 694 649 individuals of European ancestry. Hum Mol Genet 2019;28:166–174.

42. Liu M, Jiang Y, Wedow R, Li Y, Brazel DM, Chen F, et al. Association studies of up to 1.2 million individuals yield new insights into the genetic etiology of tobacco and alcohol use. Nat Genet 2019;51:237–244.

43. Oehlert GW. A Note on the Delta Method. Am Stat 1992;46:27–29.

44. Patel A, Ye T, Xue H, Lin Z, Xu S, Woolf B, et al. MendelianRandomization v0.9.0: updates to an R package for performing Mendelian randomization analyses using summarized data. Wellcome Open Res 2023;8:449.

45. Foley CN, Staley JR, Breen PG, Sun BB, Kirk PDW, Burgess S, et al. A fast and efficient colocalization algorithm for identifying shared genetic risk factors across multiple traits. Nat Commun 2021;12:764.

46. Yang J, Ferreira T, Morris AP, Medland SE, Madden PAF, Heath AC, et al. Conditional and joint multiple-SNP analysis of GWAS summary statistics identifies additional variants influencing complex traits. Nat Genet 2012;44:369–375.

47. Burgess S, Davey Smith G, Davies NM, Dudbridge F, Gill D, Glymour MM, et al. Guidelines for performing Mendelian randomization investigations: update for summer 2023. Wellcome Open Res 2023;4:186.

48. Sanderson E, Spiller W, Bowden J. Testing and correcting for weak and pleiotropic instruments in two-sample multivariable Mendelian randomization. Stat Med 2021;40:5434–5452.

49. Libby P. The changing landscape of atherosclerosis. Nature 2021;592:524–533.

50. Marston NA, Giugliano RP, Melloni GEM, Park J-G, Morrill V, Blazing MA, et al. Association of Apolipoprotein B-Containing Lipoproteins and Risk of Myocardial Infarction in Individuals With and Without Atherosclerosis: Distinguishing Between Particle Concentration, Type, and Content. JAMA Cardiol 2022;7:250–256.

51. VanderWeele TJ, Vansteelandt S. Mediation Analysis with Multiple Mediators. Epidemiol Methods 2014;2:95–115.

52. Rietveld CA, de Vlaming R, Slob EAW. The identification of mediating effects using genome-based restricted maximum likelihood estimation. PLOS Genet 2023;19:e1010638.

53. Zhou W, Bi W, Zhao Z, Dey KK, Jagadeesh KA, Karczewski KJ, et al. SAIGE-GENE+ improves the efficiency and accuracy of set-based rare variant association tests. Nat Genet 2022;54:1466–1469.

54. Chen S, Francioli LC, Goodrich JK, Collins RL, Kanai M, Wang Q, et al. A genomic mutational constraint map using variation in 76,156 human genomes. Nature 2024;625:92–100.

55. Privé F, Aschard H, Carmi S, Folkersen L, Hoggart C, O’Reilly PF, et al. Portability of 245 polygenic scores when derived from the UK Biobank and applied to 9 ancestry groups from the same cohort. Am J Hum Genet 2022;109:12–23.

56. Van Gijn ME, Ceccherini I, Shinar Y, Carbo EC, Slofstra M, Arostegui JI, et al. New workflow for classification of genetic variants’ pathogenicity applied to hereditary recurrent fevers by the International Study Group for Systemic Autoinflammatory Diseases (INSAID). J Med Genet 2018;55:530–537.

57. Cosson C, Riou R, Patoli D, Niu T, Rey A, Groslambert M, et al. Functional diversity of NLRP3 gain-of-function mutants associated with CAPS autoinflammation. J Exp Med 2024;221:e20231200.

58. Sham PC, Purcell SM. Statistical power and significance testing in large-scale genetic studies. Nat Rev Genet 2014;15:335–346.

59. The Global Cardiovascular Risk Consortium. Global Effect of Modifiable Risk Factors on Cardiovascular Disease and Mortality. N Engl J Med 2023;389:1273–1285.

60. Braddock M, Quinn A. Targeting IL-1 in inflammatory disease: new opportunities for therapeutic intervention. Nat Rev Drug Discov 2004;3:330–340.

61. Janssen CA, Oude Voshaar MAH, Vonkeman HE, Jansen TLTA, Janssen M, Kok MR, et al. Anakinra for the treatment of acute gout flares: a randomized, double-blind, placebo-controlled, active-comparator, non-inferiority trial. Rheumatology 2019;58:1344–1352.

62. Zhou X, Fernando SM, Pan AY, Laposa R, Cullen KR, Klimes-Dougan B, et al. Characterizing the NLRP3 Inflammasome in Mood Disorders: Overview, Technical Development, and Measures of Peripheral Activation in Adolescent Patients. Int J Mol Sci 2021;22:12513.

63. Libby P. Interleukin-1 Beta as a Target for Atherosclerosis Therapy: Biological Basis of CANTOS and Beyond. J Am Coll Cardiol 2017;70:2278–2289.

64. Ridker PM, MacFadyen JG, Thuren T, Libby P. Residual inflammatory risk associated with interleukin-18 and interleukin-6 after successful interleukin-1β inhibition with canakinumab: further rationale for the development of targeted anti-cytokine therapies for the treatment of atherothrombosis. Eur Heart J 2020;41:2153–2163.

65. Menu P, Pellegrin M, Aubert J-F, Bouzourene K, Tardivel A, Mazzolai L, et al. Atherosclerosis in ApoE-deficient mice progresses independently of the NLRP3 inflammasome. Cell Death Dis 2011;2:e137.

66. The Interleukin 1 Genetics Consortium. Cardiometabolic effects of genetic upregulation of the interleukin 1 receptor antagonist: a Mendelian randomisation analysis. Lancet Diabetes Endocrinol 2015;3:243–253.

67. Robinson G, Pineda-Torra I, Ciurtin C, Jury EC. Lipid metabolism in autoimmune rheumatic disease: implications for modern and conventional therapies. J Clin Invest 2022;132:e148552.

68. Vandanmagsar B, Youm Y-H, Ravussin A, Galgani JE, Stadler K, Mynatt RL, et al. The NLRP3 inflammasome instigates obesity-induced inflammation and insulin resistance. Nat Med 2011;17:179–188.

69. Ringling RE, Gastecki ML, Woodford ML, Lum-Naihe KJ, Grant RW, Pulakat L, et al. Loss of Nlrp3 Does Not Protect Mice from Western Diet-Induced Adipose Tissue Inflammation and Glucose Intolerance. PLOS ONE 2016;11:e0161939.

70. Pierantonelli I, Rychlicki C, Agostinelli L, Giordano DM, Gaggini M, Fraumene C, et al. Lack of NLRP3-inflammasome leads to gut-liver axis derangement, gut dysbiosis and a worsened phenotype in a mouse model of NAFLD. Sci Rep 2017;7:12200.

71. Henao-Mejia J, Elinav E, Jin C, Hao L, Mehal WZ, Strowig T, et al. Inflammasome-mediated dysbiosis regulates progression of NAFLD and obesity. Nature 2012;482:179–185.

72. Netea MG, Joosten LAB, Lewis E, Jensen DR, Voshol PJ, Kullberg BJ, et al. Deficiency of interleukin-18 in mice leads to hyperphagia, obesity and insulin resistance. Nat Med 2006;12:650–656.

73. Krishnan SM, Ling YH, Huuskes BM, Ferens DM, Saini N, Chan CT, et al. Pharmacological inhibition of the NLRP3 inflammasome reduces blood pressure, renal damage, and dysfunction in salt-sensitive hypertension. Cardiovasc Res 2019;115:776–787.

74. Amancherla K, Wells JA, Bick AG. Clonal hematopoiesis and vascular disease. Semin Immunopathol 2022;44:303–308.

75. Svensson EC, Madar A, Campbell CD, He Y, Sultan M, Healey ML, et al. TET2-Driven Clonal Hematopoiesis and Response to Canakinumab: An Exploratory Analysis of the CANTOS Randomized Clinical Trial. JAMA Cardiol 2022;7:521–528.

76. Martinon F, Pétrilli V, Mayor A, Tardivel A, Tschopp J. Gout-associated uric acid crystals activate the NALP3 inflammasome. Nature 2006;440:237–241.

77. Klein AL, Imazio M, Cremer P, Brucato A, Abbate A, Fang F, et al. Phase 3 Trial of Interleukin-1 Trap Rilonacept in Recurrent Pericarditis. N Engl J Med 2021;384:31–41.

78. Senkevich K, Liu L, Alvarado CX, Leonard HL, Nalls MA, Global Parkinson’s Genetics Program (GP2), et al. Lack of genetic evidence for NLRP3 inflammasome involvement in Parkinson’s disease pathogenesis. NPJ Park Dis 2024;10:145.

79. Gill D, Dib M-J, Cronjé HT, Karhunen V, Woolf B, Gagnon E, et al. Common pitfalls in drug target Mendelian randomization and how to avoid them. BMC Med 2024;22:473.

80. Burgess S, Davies NM, Thompson SG. Bias due to participant overlap in two-sample Mendelian randomization. Genet Epidemiol 2016;40:597–608.

81. Davey Smith G, Timpson N, Ebrahim S. Strengthening causal inference in cardiovascular epidemiology through Mendelian randomization. Ann Med 2008;40:524–541.

82. Chang CC, Chow CC, Tellier LC, Vattikuti S, Purcell SM, Lee JJ. Second-generation PLINK: rising to the challenge of larger and richer datasets. GigaScience 2015;4:7.

83. Zhu X, Feng T, Tayo BO, Liang J, Young JH, Franceschini N, et al. Meta-analysis of Correlated Traits via Summary Statistics from GWASs with an Application in Hypertension. Am J Hum Genet 2015;96:21–36.

84. Ritchie SC, Surendran P, Karthikeyan S, Lambert SA, Bolton T, Pennells L, et al. Quality control and removal of technical variation of NMR metabolic biomarker data in ∼120,000 UK Biobank participants. Sci Data 2023;10:64.

85. Astle WJ, Elding H, Jiang T, Allen D, Ruklisa D, Mann AL, et al. The Allelic Landscape of Human Blood Cell Trait Variation and Links to Common Complex Disease. Cell 2016;167:1415–1429.e19.

86. Wu T, Hu E, Xu S, Chen M, Guo P, Dai Z, et al. clusterProfiler 4.0: A universal enrichment tool for interpreting omics data. The Innovation 2021;2:100141.

87. Dolgalev I. msigdbr: MSigDB Gene Sets for Multiple Organisms in a Tidy Data Format. 2025. 10.32614/CRAN.package.msigdbr.

88. Gene Ontology Consortium. The Gene Ontology knowledgebase in 2026. Nucleic Acids Res 2026;54:D1779–D1792.

89. Benjamini Y, Hochberg Y. Controlling the False Discovery Rate: A Practical and Powerful Approach to Multiple Testing. J R Stat Soc Ser B Methodol 1995;57:289–300.

90. Mangan MSJ, Olhava EJ, Roush WR, Seidel HM, Glick GD, Latz E. Targeting the NLRP3 inflammasome in inflammatory diseases. Nat Rev Drug Discov 2018;17:588–606.

